# Probiotics and the EARly Life effects on intestinal bacteria and inflammation in children with Cystic Fibrosis (The “PEARL-CF” Study)

**DOI:** 10.64898/2025.12.04.25341620

**Authors:** Michael J Coffey, Josie van Dorst, B.L.D. Uthpala Pushpakumara, Jessica Halim, Jennifer Hudson, Leanne Plush, Katrina Christi, Charles Foster, Rosemary Carzino, Natalie Zajakovski, Laura Appleton, Joyce Cheney, Judy Wood, Sarah Jane Monaghan, William Rawlinson, Andrew S Day, Mark Oliver, Sarath Ranganathan, Hiran Selvadurai, Claire Wainwright, Torsten Thomas, Adam Jaffe, Chee Y Ooi

## Abstract

The role of probiotics in children with cystic fibrosis (CwCF) remains unclear. The PEARL-CF study was an international, double-blind, randomized, placebo-controlled study involving CwCF (0–6 years), and assessed the effects of a probiotic on intestinal microbiota and clinical outcomes. A multi-strain probiotic (15 Lactobacillus/Bifidobacterium strains; ∼2-3×10^10^ CFU daily) or placebo was administered for 12-months and participants followed a further 12-months post-intervention. Among 77 CwCF (38 probiotic, 39 placebo), bacterial alpha diversity did not differ between groups. Participants ≥4 years on probiotic demonstrated higher bacterial richness (post-hoc analysis). *Haemophilus influenzae* was less prevalent in clinically indicated respiratory swabs in the probiotic cohort during the intervention (27% vs 56%; p=0.03), especially when commenced prior to 4 years old. Fecal M2-pyruvate kinase decreased in the probiotic cohort from baseline to 12 months. This study provides support for multi-strain probiotics in CwCF, even in the era of modulator therapy.

## Introduction

Cystic fibrosis (CF) is a life-limiting genetic disease caused by mutations in the cystic fibrosis transmembrane conductance regulator (*CFTR*) gene affecting over 160,000 children and adults worldwide^1^. It is a multi-system disease predominantly affecting the lungs, gastrointestinal tract, pancreas and liver. Chronic and recurrent respiratory infections are the leading cause of reduced life expectancy and increased morbidity in individuals with CF^2^. Growth and nutritional status of people with CF is paramount, as they are major determinants of lung function and overall survival^3,4^.

The gastrointestinal microbiota play an important role in health and disease, contributing to metabolic function, immunity and host nutrition^5^. The intestinal microbiome may also influence susceptibility to respiratory infections in CF, highlighting the importance of the gut-lung axis^6^. In healthy populations, the gastrointestinal microbiota develop rapidly during the first year of life and by 3-4 years of age the composition and diversity resembles that of an adult^7^. Once established, the microbiota is relatively stable and difficult to perturb^7,8^ In children with CF (CwCF), there is a persistent delay in the maturation of the gut microbiota beyond the first 3 years of life, furthermore there is a shift towards an increase in potentially pathogenic species and a reduction in commensals^9-14^. Disruptions to intestinal microbiota during this “critical window” may have lasting health implications.^15^ Gastrointestinal dysbiosis is well documented in people with CF and hypothesized to be due to the dehydrated, acidic luminal environment, inspissated slow-to-clear mucus and low-grade inflammation within the gut^16^. Fecal microbial and metabolomic factors, including bile acids and microbial metabolites may impact nutrition, growth, and GI health in infants with CF^17^. Furthermore, poor growth is associated with intestinal inflammation in CwCF^18^.

Probiotics are live microorganisms, which when consumed in adequate amounts confer a health benefit on a host^19^. Probiotics may be beneficial in several pediatric conditions, with the best efficacy seen in the treatment of infectious gastroenteritis, and prevention of antibiotic-associated, nosocomial and *Clostridioides difficile*-associated diarrhea.^20^ A recent Cochrane Review of probiotics for people with CF demonstrated that probiotics reduce fecal calprotectin in children and adults^21^. To date it remains unclear if probiotics have a benefit when used prior to the establishment of the intestinal microbiota. Additionally, it is unknown if any benefits persist once supplementation is ceased.

The <u>P</u>robiotics and the <u>EAR</u>ly <u>L</u>ife effects on intestinal bacteria and inflammation in children with <u>C</u>ystic <u>F</u>ibrosis Study (The “PEARL-CF” Study) aimed to evaluate the extent of the effects of probiotics on the intestinal microbiota, intestinal inflammation and clinical outcomes in CwCF. It also aimed to evaluate whether probiotics restore “normality” by including a group of non-CF healthy controls (HC), matched for age and gender. Furthermore, this study aimed to identify a potential “therapeutic window” of intervention before the maturation of intestinal microbiota composition. This study also aimed to assess for any prolonged effects once supplementation was ceased.

The hypothesis for this study was that probiotics would restore the abnormal gut microbiota in CwCF, which in turn reduces intestinal inflammation. Additional hypotheses included: (i) probiotics will have clinical benefits for CwCF, and (ii) when probiotics are given in early life, the effects of probiotics, even when ceased, are sustained compared to when probiotics are given after the gut microbiota has become established.

## Results

### Population characteristics

Between June 2017 to December 2022, 77 CwCF were recruited. Thirty-eight and 39 CwCF were randomized to receive the probiotic or placebo (Extended Data Table 1), respectively (Fig. 1). An additional 70 HC were recruited. Overall, the two cohorts of CwCF were balanced with respect to baseline characteristics (Table 1). Prior to commencement, one (3%) child in the probiotic and five (13%) in the placebo group withdrew. In the probiotic cohort, 26 (68%) and 23 (61%) participants completed the intervention and post-intervention periods, respectively. In the placebo cohort, 23 (59%) and 21 (54%) participants completed the intervention and post-intervention periods, respectively.

**Figure 1.**
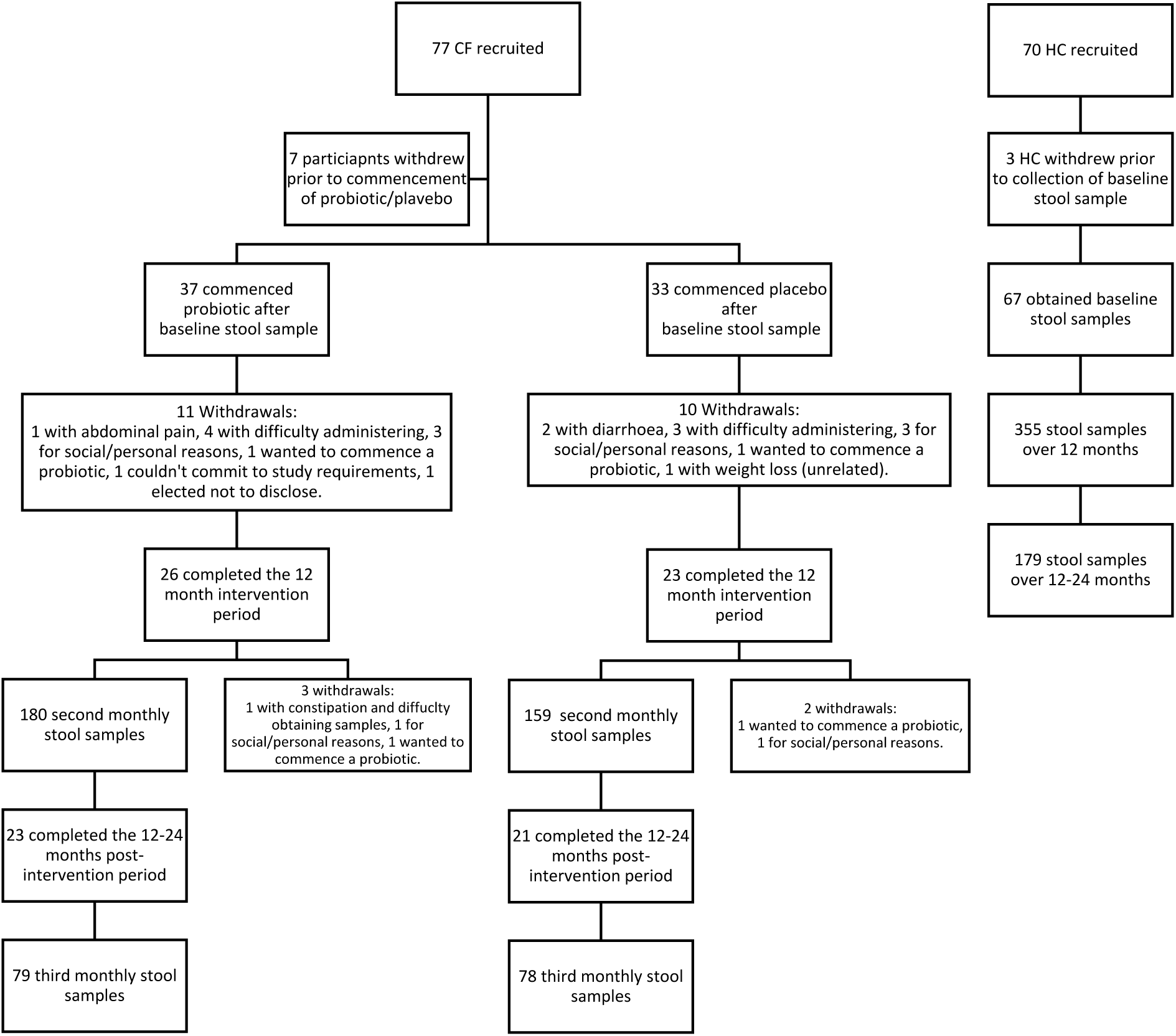
Study CONSORT diagram participants and samples.

**Table 1.** Baseline characteristics of CF participants.

|  | **Probiotic Cohort**<br>***n* = 38** | **Placebo Cohort**<br>***n* = 39** | **P-value** |
| --- | --- | --- | --- |
| **Age, mean years (*SD*)** | 3.2 (1.6) | 3.1 (1.7) | 0.7 |
| Under 3 years, *n* | 18 | 20 |  |
| Over 3 years, *n* | 20 | 19 |  |
| **Male sex, *n* (%)** | 22 (58%) | 23 (59%) | 0.99 |
| **Pancreatic insufficiency, *n* (%)** | 33 (87%) | 35 (90%) | 0.7 |
| **Genetic mutations** |  |  | 0.8 |
| *F508del* / *F508del*, *n* (%) | 15 (39%) | 12 (31%) |  |
| *F508del* / Other, *n* (%) | 14 (37%) | 14 (36%) |  |
| Other, *n* (%) | 2 (5%) | 3 (8%) |  |
| Not reported, *n* (%) | 7 (18%) | 10 (26%) |  |
| **Participant's Site** |  |  | 0.8 |
| Sydney Children's Hospital, *n* | 9 | 8 |  |
| Royal Children's Hospital, *n* | 18 | 17 |  |
| Christchurch Hospital, *n* | 0 | 2 |  |
| Children's Hospital at Westmead, *n* | 8 | 8 |  |
| Queensland Children's Hospital, *n* | 3 | 4 |  |
| **Self-reported ethnicity\*** |  |  | 0.3 |
| Australian Aboriginal & Torres Strait Islander, *n* | 0 | 5 |  |
| European, *n* | 6 | 4 |  |
| New Zealand Maori, *n* | 0 | 1 |  |
| Oceanian, *n* | 28 | 24 |  |
| Other, *n* | 3 | 0 |  |
| Not reported, *n* | 6 | 9 |  |
| **Anthropometrics** |  |  |  |
| Weight z-score, mean (*SD*) | 0.3 (0.9) | 0.1 (0.9) | 0.4 |
| Height z-score, mean (*SD*) | 0.02 (0.9) | -0.08 (0.9) | 0.7 |
| **Medications** |  |  |  |
| Ivacaftor, *n* (%) | 4 (11%) | 1 (3%) | 0.2 |
| PERT, *n* (%) | 33 (87%) | 35 (90%) | 0.7 |
| PPI, *n* (%) | 30 (79%) | 30 (77%) | 0.3 |
| Prophylactic Antibiotic(s), *n* (%) | 26 (68%) | 18 (46%) | 0.2 |

\* Participants can report multiple ethnicities. PERT, pancreatic enzyme replacement therapy. PPI, proton pump inhibitor.

### Primary outcomes

Fecal samples were analyzed for bacterial alpha diversity (Fig. 2 and Table 2). Fecal bacterial richness from all samples across both the intervention and post-intervention periods illustrated the differences in alpha diversity between CwCF and HC across all ages (Fig. 2A).

**Figure 2.**
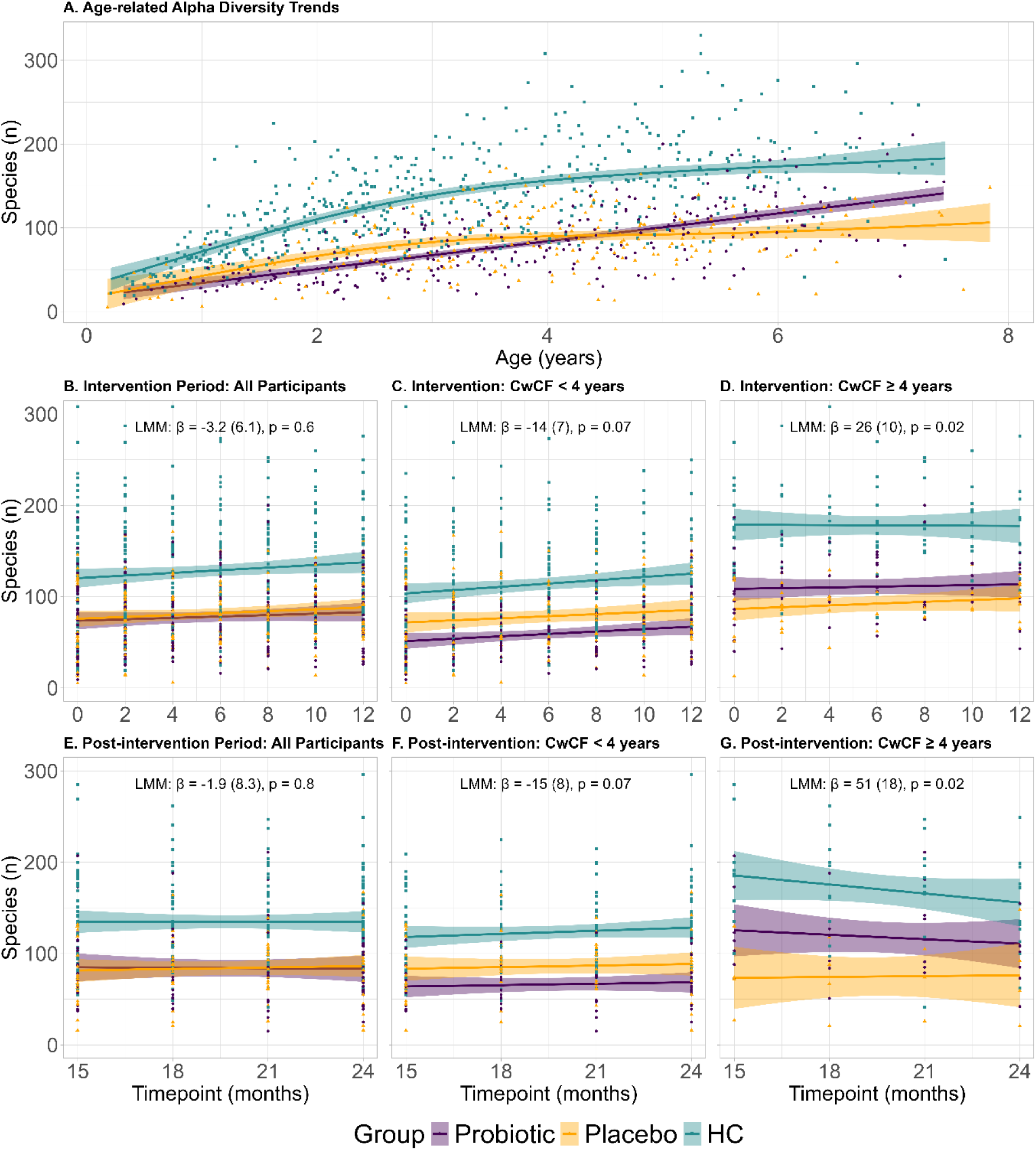
Fecal bacterial alpha diversity (observed species richness). Scatterplots of fecal bacterial species richness. Diversity trajectories were modelled using generalized additive models with the estimated group mean trend (lines) and 95% confidence intervals (shaded regions). **Fig. A** presents the age-related alpha diversity trends for all samples at all timepoints (baseline to 24 months). **Fig. B** and **E** present the results for all participants during the intervention period. **Fig. C** and **F** present participants who commenced the probiotic/placebo < 4 years of age, and **Fig. D** and **G** present participants who commenced the probiotic/placebo ≥ 4 years of age (post hoc analysis). For **Fig. B-G**, linear mixed-effects models (LMM) were used to quantify the longitudinal change in species richness whilst accounting for intra-individual repeated measured. The LMM results are presented as the model-estimated slope (β coefficient), standard error (SE), and p-value. Significance was defined as *p* < 0.05.

**Table 2.** Fecal alpha diversity indices & inflammatory markers at baseline, and during the intervention and post-interventions periods.

| Diversity Indices | Probiotic Cohort<br>*n* = 37 | Placebo Cohort<br>*n* = 33 | LMM Estimate (SE)* | P-value | Healthy Controls<br>*n*=69 |
| --- | --- | --- | --- | --- | --- |
| **A. Richness, *n* OTUs** |  |  |  |  |  |
| Baseline | 69 (35 – 106) | 70 (45 – 118) |  | 0.7 | 112 (62 – 166) |
| 12 months | 92 (51 – 114) | 83 (67 – 114) |  | 0.6 | 126 (98 – 174) |
| Intervention |  |  | -3.2 (6.1) | 0.6 |  |
| 24 months | 73 (59 – 108) | 86 (62 – 98) |  | 0.7 | 129 (100 – 161) |
| Post-intervention |  |  | -1.9 (8.3) | 0.8 |  |
| **B. Shannon Diversity Index** |  |  |  |  |  |
| Baseline | 2.4 (0.8) | 2.5 (0.8) |  | 0.7 | 2.8 (0.9) |
| 12 months | 2.6 (0.7) | 2.7 (0.5) |  | 0.3 | 3.1 (0.6) |
| Intervention |  |  | -0.05 (0.1) | 0.6 |  |
| 24 months | 2.5 (0.6) | 2.6 (0.8) |  | 0.7 | 3.1 (0.5) |
| Post-intervention |  |  | -0.07 (0.1) | 0.6 |  |
| **C. M2PK, U/mL** |  |  |  |  |  |
| Baseline | 6.6 (2.3 – 12.5) | 5.4 (2.5 – 10.4) |  | 0.5 | 3.3 (1.2 – 7.5) |
| 12 months | 4.3 (2.2 – 8.5) | 11.3 (2.5 – 19.6) |  | 0.053 | 1.5 (0.8 – 2.4) |
| Delta baseline to 12 months | -1.3 (-4.3 – 1.1) | 3.9 (-1.0 – 11.9) |  | 0.002 |  |
| Intervention |  |  | -3.6 (5.5) | 0.5 |  |
| 24 months | 5.2 (3.2 – 15.7) | 7.4 (2.9 – 20.4) |  | 0.9 | 1.9 (0.7 – 3.1) |
| Post-intervention |  |  | 2.2 (9.2) | 0.8 |  |
| **D. Calprotectin, mg/kg** |  |  |  |  |  |
| Baseline | 144 (50 – 283) | 140 (48 – 274) |  | 0.8 | 64 (34 – 108) |
| 12 months | 157 (83 – 301) | 129 (59 – 360) |  | 0.8 | 52 (28 – 114) |
| Intervention |  |  | 10 (57) | 0.9 |  |
| 24 months | 123 (67 – 236) | 98 (51 – 275) |  | 0.7 | 37 (20 – 79) |
| Post-intervention |  |  | 19 (71) | 0.8 |  |

Data presented as mean (SD), median (IQR) or estimate (SE). \*Probiotic cohort is the reference group for the linear mixed model (LMM) estimates with standard error (SE), i.e. a positive value would indicate the result is higher in the probiotic cohort compared with the placebo cohort; OTUs, operational taxonomic units

Fecal alpha diversity assessed using species richness and Shannon diversity index did not differ between the probiotic and placebo groups at baseline (Table 2A-B). During the intervention period, alpha diversity indices did not differ between the probiotic and placebo groups: (i) species richness linear mixed model (LMM) estimate, standard error (SE) was 3.2 (6.1) species lower in the probiotic group, p = 0.6 (Fig. 2B and Table 2A), and (ii) Shannon index LMM, (SE) – 0.05 (00.1) lower in the probiotic group, p = 0.6 (Table 2B). Similarly, alpha diversity indices did not differ between the probiotic and placebo groups during the post-intervention period (Fig. 2E and Table 2A-B).

Fecal beta diversity was assessed using Aitchison distance (Fig. 3A-D) as it better handled the data compositionality compared to Bray-Curtis dissimilarity (Extended Data Figure 2) Beta diversity did not differ between the probiotic and placebo groups at baseline (PERMANOVA: R² = 0.014, p = 0.5; Fig. 3A), 12 months (R² = 0.020, p = 0.8; Fig. 3B), or 24 months (R² = 0.025, p = 0.3; Fig. 3C). Longitudinal analysis of beta diversity between the probiotic and placebo groups during the intervention period showed a small overall difference (F = 5.98, p = 0.017; Fig. 3D). Time was the strongest factor affecting beta diversity in CwCF (LMM: F = 356, p < 2×10⁻¹⁶; Fig. 3D).

**Figure 3.**
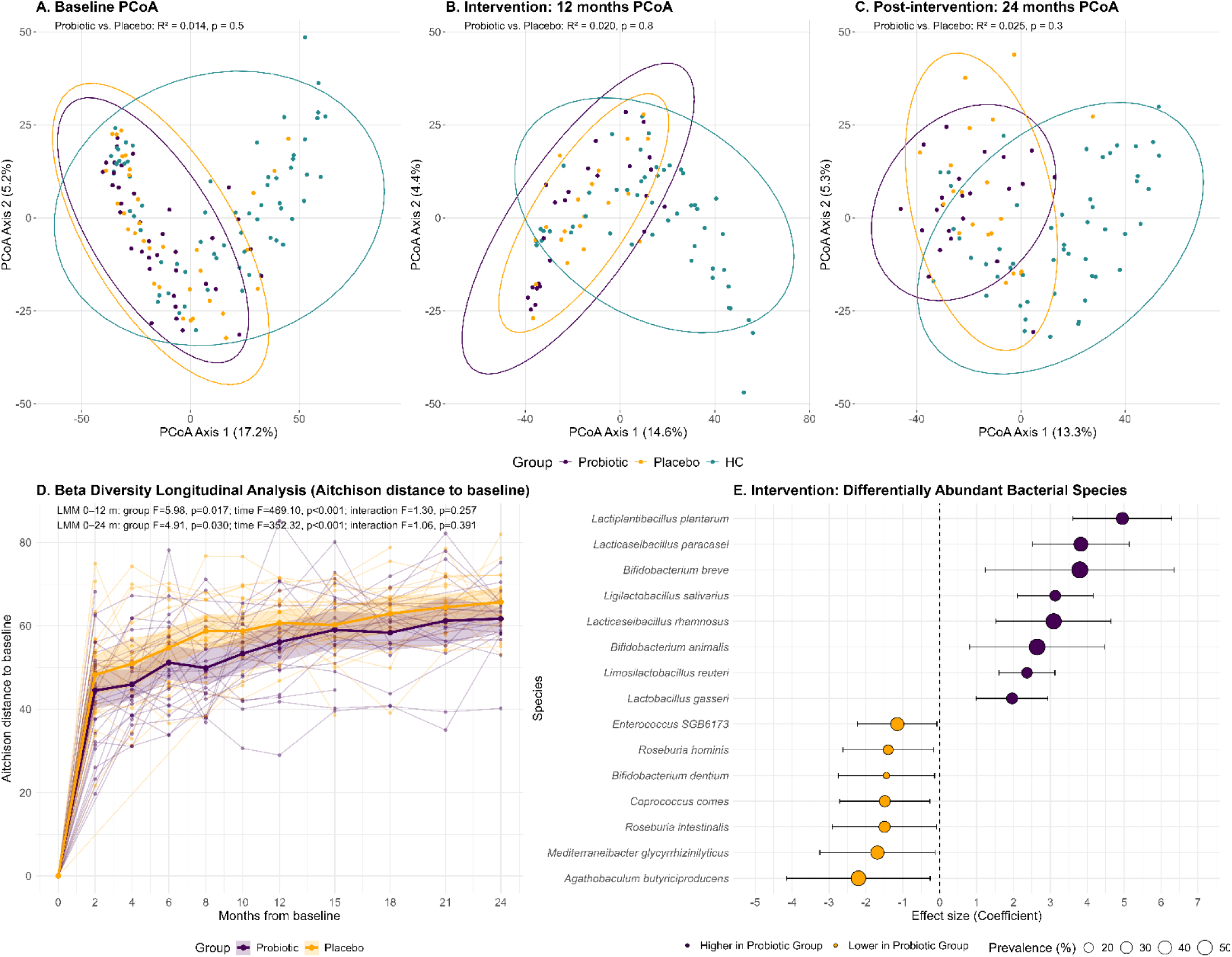
Fecal bacterial beta diversity (Aitchison distance) and differentially abundant fecal bacterial species (during the intervention period). Principal Coordinates Analysis (PCoA) of Aitchison distance based on CLR-transformed species abundances (with 95% confidence ellipses) at baseline (**A**), end of the 12 month intervention period (**B**), and end of the 24 month post-intervention period (**C**). In graphs A-C, PERMANOVA results are for probiotic vs. placebo analyses (not including healthy controls). **Fig. D** presents a longitudinal analysis of beta diversity trends from baseline to 24 months for all CwCF. Lines show individual trajectories with overlaid group means and 95% confidence intervals. Linear mixed-effects models were used to analyse treatment group and time effects during the intervention period (0-12 m) and the entire study period (0-24 m). **Fig. E** presents the differentially abundant fecal bacterial species during the intervention period from CwCF on the probiotic and placebo. Effect sizes (coefficients with 95% CI) are plotted for species significantly associated with treatment group (*q* < 0.25). Positive coefficients indicate higher abundance in the probiotic group; negative values indicate higher abundance in the placebo group.

There was evidence that the bacteria of the probiotic formulation (Extended Data Table 1) colonized the gut of CwCF (Fig. 3E and Extended Data Table 2). The relative abundance of eight of the fifteen probiotic strains was increased in the fecal samples from the probiotic cohort compared to the placebo cohort during the intervention period (Fig. 3E; q < 0.25 for all). Probiotic supplementation was associated with a relative reduction in several bacterial species during the intervention period (Fig. 3E; q < 0.25 for all). During the post-intervention period, no bacterial species were differentially abundant between the probiotic and placebo cohorts (q > 0.25 for all).

### Secondary outcomes

Clinical outcomes were assessed over the intervention period (Table 3). During the 12-month intervention period, participants underwent respiratory pathogen testing only if clinically indicated for a pulmonary infection (Table 3A). During the intervention period, fewer participants in the probiotic cohort had a positive *Haemophilus influenzae* result compared to the placebo cohort (27% vs 58%, respectively, p = 0.02). Additionally, fewer participants in the probiotic cohort had their first-ever positive *H. influenzae* result compared to the placebo cohort (16% vs. 48%, respectively, p = 0.008). Rates of hospitalizations, pulmonary exacerbations and therapeutic antibiotic courses did not significantly differ between the probiotic and placebo cohorts during the intervention period (Table 3B-C).

**Table 3.** Clinical outcomes during the 12-month intervention period.

| Clinical Outcome | Probiotic Cohort<br>*n* = 37 | Placebo Cohort<br>*n* = 33 | LMM Estimate<br>(SE)* | P-value |
| --- | --- | --- | --- | --- |
| **A. Participants with CF-associated respiratory pathogens isolated from clinical samples** <sup>†</sup> |  |  |  |  |
| *Haemophilus influenzae*, *n* (%) | 10 (27%) | 19 (58%) |  | 0.02 |
| *Staphylococcus aureus* | 14 (38%) | 11 (33%) |  | 0.9 |
| Chronic *S. auerus* | 5 (14%) | 1 (3%) |  | 0.3 |
| Non-mucoid *Pseudomonas aeruginosa* | 3 (8%) | 5 (15%) |  | 0.6 |
| *Stenotrophomonas spp.* | 2 (5%) | 2 (6%) |  | 0.99 |
| *Klebsiella spp.* | 1 (3%) | 0 |  | 0.99 |
| *Acinetobacter spp.* | 0 | 3 (9%) |  | 0.2 |
| **B. Hospitalizations for a pulmonary exacerbation** |  |  |  |  |
| Participants with at least 1 hospitalization, *n* (%) | 6 (16%) | 9 (27%) |  | 0.4 |
| Time to first hospitalization, median months (IQR) | 5 (4 – 6) | 2 (2 – 4) |  | 0.2 |
| Number of hospitalizations, median *n* (IQR) | 0 (0 – 0) | 0 (0 – 1) |  | 0.3 |
| **C. Therapeutic antibiotic courses (oral or inhaled)** |  |  |  |  |
| Participants with at least 1 course, *n* (%) | 29 (78%) | 26 (79%) |  | 0.99 |
| Time to first antibiotic, median months (IQR) | 2 (2 – 4) | 2 (2 – 4) |  | 0.8 |
| Number of courses | 2 (1 – 2) | 2 (1 – 3) |  | 0.96 |
| **D. Anthropometrics** |  |  |  |  |
| **Weight z-score** |  |  |  |  |
| Baseline | 0.3 (0.9) | 0.2 (0.9) |  | 0.5 |
| 12 months | 0.5 (0.9) | 0.3 (0.9) |  | 0.3 |
| Intervention |  |  | 0.1 (0.2) | 0.7 |
| **Height z-score** |  |  |  |  |
| Baseline | 0.02 (0.9) | -0.06 (0.9) |  | 0.7 |
| 12 months | 0.5 (0.9) | 0.1 (1.3) |  | 0.3 |
| Intervention |  |  | 0.2 (0.2) | 0.4 |
| **Body Mass Index z-score (≥ 2 years old)** |  |  |  |  |
| Number (% total) | 28 (76%) | 24 (73%) |  |  |
| Baseline | 0.7 (0.6) | 0.4 (0.8) |  | 0.1 |
| 12 months | 0.2 (0.8) | 0.6 (0.6) |  | 0.1 |
| Intervention |  |  | 0.09 (0.2) | 0.7 |
| **E. Health-related Quality of Life** |  |  |  |  |
| **Baseline Survey Completed** | 36 (97%) | 32 (97%) |  |  |
| PedsQL Infant Scales (ages 1-12 months), n (% total) | 6 (17%) | 4 (13%) |  |  |
| PedsQL Infant Scales (ages 13-24 months), n (% total) | 6 (17%) | 5 (16%) |  |  |
| PedsQL GI Symptoms (ages 2-4 years), n (% total) | 19 (53%) | 19 (59%) |  |  |
| PedsQL GI Symptoms (ages 5-7 years), n (% total) | 5 (14%) | 4 (13%) |  |  |
| **PedsQL Total Score** |  |  |  |  |
| Baseline PedsQL Total Score<sup>‡</sup> | 85 (74 – 92) | 83 (75 – 93) |  | 0.8 |
| 12 months PedsQL Total Score<sup>‡</sup> | 93 (82 – 99) | 90 (79 – 91) |  | 0.3 |
| Intervention PedsQL Total Score<sup>‡</sup> |  |  | 0.7 (2.8) | 0.8 |

\*Probiotic cohort is the reference group for the linear mixed model (LMM) estimates with standard error (SE), i.e. a positive value would indicate the result is higher in the probiotic cohort compared with the placebo cohort. <sup>†</sup>Microbiology = intention to treat analysis of intervention period (swab, sputum or bronchiolar lavage samples). <sup>‡</sup>PedsQL Total Score, reported out of 100, higher indicates better quality of life; age-appropriate survey results combined for the overall analysis.

Regarding anthropometric data, there was no baseline difference in weight, height or body mass index (BMI) z-scores between the probiotic and placebo cohorts (Table 3D), and the mean z-scores suggest the current cohort were well grown at baseline. There was no overall difference during the intervention period in weight, height or body mass index (BMI) z-scores between the probiotic and placebo cohorts (p>0.05 for all).

Intestinal inflammation was measured using fecal M2-pyruvate kinase (M2PK) and calprotectin (Table 2C-D; Extended Data Fig. 2-3). There was no baseline difference in M2PK between the probiotic and placebo groups. Fecal M2PK at the end of the intervention was lower in the probiotic cohort compared to the placebo cohort and was close to the cut-off for significance (4.3 U/mL (2.2 – 8.5) vs. 11.3 (2.5 – 19.6), respectively, p = 0.053). The change in fecal M2PK from baseline to 12 months was lower in the probiotic cohort compared with placebo cohort (-1.3 (-4.3 – 1.1) vs. 3.9 (-1.0 – 11.9), respectively, p=0.002). There was no overall significant difference in M2PK between the probiotic and placebo groups when examining the entire intervention period (LMM estimate (SE) 3.6 U/mL (5.5) lower in the probiotic cohort, p = 0.5; Extended Data Figure 2A/D). During the post-intervention period, fecal M2PK did not significantly differ between the probiotic and placebo groups (Extended Data Figure 2G/J). Fecal calprotectin did not significantly differ between the probiotic and placebo groups at baseline, during the intervention or post-intervention periods (Table 2D; Extended Data Figure 3A/D and 3G/J).

Health-related quality of life (HRQOL) was assessed using age-appropriate PedsQL surveys (Table 4E). Overall HRQOL did not differ between the probiotic and placebo groups at baseline or during the intervention period (p>0.05 for all).

### Tertiary outcomes

Lung function testing was limited during the COVID-19 pandemic and no meaningful comparisons could be made between the probiotic and placebo groups. Numbers of lung function tests performed are presented in Extended Data Table 3.

### Safety outcomes

There were no serious adverse events reported during the study. A single participant in the probiotic cohort reported abdominal pain and withdrew from the study. A single participant in the probiotic cohort experienced diarrhea 2 days after probiotic commencement, however, was able to restart the probiotic 1 week later without any issues. Two participants in the placebo cohort experienced diarrhea and withdrew from the study.

### Sensitivity outcomes

The fecal alpha diversity, beta diversity, M2PK and calprotectin results presented above were analyzed using a modified intention-to-treat approach, with a last result carried forward to handle missing data (described in Methods section). A sensitivity per-protocol analysis was also performed and demonstrated no relevant changes to the results of the intention-to-treat analysis.

A sensitivity analysis of prophylactic antibiotics, any antibiotic use, proton pump inhibitors (PPI), pancreatic enzyme replacement therapy (PERT) and ivacaftor was performed by including them as a random effect in the linear mixed-effects models on alpha diversity. Both prophylactic antibiotics (β = –8.8 ± 4.3 SE, *p* = 0.043; Extended Data Fig. 4) and any antibiotic use (β = –15.6 ± 4.3 SE, *p* < 0.001; Extended Data Fig. 5) were associated with lower richness, independent of the probiotic/placebo group and age. In contrast, no significant associations were observed for PPI therapy (β = –4.5 ± 5.7 SE, *p* = 0.43; Extended Data Fig. 6), PERT use (β = –1.7 ± 4.2 SE, *p* = 0.69; Extended Data Fig. 7), or ivacaftor therapy (β = 0.27 ± 6.7 SE, *p* = 0.97; Extended Data Fig. 8). Longitudinal patterns remained consistent across all medication strata providing support that the primary outcome results were not significantly influenced by concomitant medication use.

### Post hoc analyses

Given the community structure of the gut microbiota is highly resilient and relatively stable once it reaches an adult-like state^7,8,15^, this study aimed to determine if there was a therapeutic window in early childhood. A blinded analysis of fecal alpha diversity (observed richness) was performed and identified a clear inflection point and plateau in alpha diversity around 4 years of age (Extended Data Fig. 9). Based on this result and prior to unblinding, it was elected to stratify the CF cohorts into < 4 years of age and ≥ 4 years of age at commencement of the probiotic or placebo supplement.

#### Children aged < 4 years at study commencement

Regarding CwCF aged < 4 years at study start, 22 and 24 were randomized to the probiotic and placebo groups, respectively. During the intervention period, CwCF in the probiotic cohort had fewer pulmonary pathogen test results positive with *H. influenzae* compared to the placebo cohort, (18% vs 62%, respectively, p=0.0006). Fecal bacterial richness for the probiotic and placebo cohorts during the intervention period showed a nonsignificant trend towards lower richness in the probiotic cohort (estimate (SE) -14 (7) species, p = 0.07; Fig. 2C). Fecal beta diversity did not differ between the probiotic and placebo groups at baseline (PERMANOVA: R² = 0.028, p = 0.12; Extended Data Fig. 10A), 12 months (R² = 0.032, p = 0.45; Extended Data Fig. 10B), or 24 months (R² = 0.041, p = 0.058; Extended Data Fig. 10C). Longitudinal analysis of beta diversity between the probiotic and placebo groups during the intervention period showed a marked difference in the progression of the microbial composition (F = 13.14, p = 0.0007; Extended Data Fig. 10D).

There was evidence for bacteria of the probiotic formulation (Extended Data Table 1) to colonize the gut of CwCF < 4 years at study commencement (Extended Data Fig. 10E and Extended Data Table 4). Eight of the 15 probiotic strains were increased in the fecal samples from the probiotic cohort compared to the placebo cohort during the intervention period. Probiotic supplementation was associated with a relative reduction in 25 bacterial species during the intervention period (Extended Data Table 4). Further, in the post-intervention period, probiotic use was associated with the following relative changes: (i) *C. difficile* was increased, and (ii) *Faecalibacterium prausnitzii* was decreased in the probiotic cohort compared to the placebo cohort (Extended Data Table 4).

Notably, the relative abundance of *Agathobaculum butyriciproducens* (a butyrate producer) was lower in the probiotic cohort compared to the placebo cohort during the intervention period (coefficient (SE) -2.57 (0.68), q = 0.012; Extended Data Figure 11A). Targeted metabolomics identified lower levels of fecal butyric acid in the probiotic cohort compared to the placebo cohort during the intervention period (estimate (SE) 2125 μg/g (818) dry weight, p = 0.01; Extended Data Figure 11B). *A. butyriciproducens* remained lower in the probiotic cohort compared to the placebo cohort during the post-intervention period (estimate (SE) 0.24% (0.11), p = 0.04; Extended Data Figure 12A). At 24 months (the end of the post-intervention period), fecal butyric acid levels also remained lower in the probiotic cohort compared to the placebo cohort (1853 μg/g (1155 – 3389) vs. 5439 μg/g (2661 – 11704), respectively, p = 0.01; Extended Data Figure 12B).

#### Children aged ≥ 4 years at study commencement

Regarding CwCF aged ≥ 4 years at study start, 15 and nine were randomized to the probiotic and placebo groups, respectively. Fecal bacterial richness was higher in the probiotic cohort than the placebo cohort during the intervention period (estimate (SE) 26 (10) species, p = 0.02); Extended Data Figure 5B) and post-intervention period (estimate (SE) 51 (18) species, p = 0.02; Extended Data Figure 6B). Fecal beta diversity did not differ between the probiotic and placebo groups at baseline (PERMANOVA: R² = 0.039, p = 0.82; Extended Data Fig. 13A), 12 months (R² = 0.086, p = 0.57; Extended Data Fig. 13B), or 24 months (R² = 0.095, p = 0.058; Extended Data Fig. 13C). Longitudinal analysis of beta diversity between the probiotic and placebo groups during the intervention period showed no difference in the progression of the microbial composition (F = 0.16, p = 0.7; Extended Data Fig. 13D).

In CwCF who commenced the probiotic ≥ 4 years of age, there was evidence for bacteria to colonize within the gut (Extended Data Fig. 13E and Extended Data Table 4), albeit to a lesser extent than for CwCF starting the probiotic < 4 years of age. Four of the 15 probiotic strains were increased in the fecal samples from the probiotic cohort compared to the placebo cohort during the intervention period (Extended Data Table 4). Probiotic supplementation was not associated with a relative increase or decrease in other bacterial strains during the intervention period. There was also no evidence of a sustained effect after probiotic supplementation (Extended Data Table 4).

#### Fecal butyric acid levels in children with cystic fibrosis (all ages)

Fecal butyric acid levels trended towards being lower in the probiotic cohort compared to the placebo cohort during the intervention period (estimate (SE) 1443 μg/g (741) dry weight, p = 0.057; Extended Data Figure 14A). Fecal butyric acid levels were lower in the probiotic cohort compared to the placebo cohort at the end of the post-intervention period (24 months), 1984 μg/g (1283 – 4244) vs. 5283 μg/g (2535 – 11154), respectively, p = 0.04 (Extended Data Figure 14B).

## Discussion

The PEARL-CF study provides the most comprehensive assessment to date of the efficacy of a multi-strain probiotic supplement for CwCF aged 0-6 years. This study identified that in young CwCF, a multi-strain probiotic (predominantly *Lactobacillus* and *Bifidobacterium* strains) was associated with a reduced rate of *H. influenzae* detection in clinically indicated respiratory pathogen tests, particularly if the probiotic commenced prior to 4 years of age. The PEARL-CF study also provides proof-of-concept that there is a therapeutic window (prior to 4 years of age) during which modulating the gut microbiota can have sustained effects well beyond cessation of the probiotic. Despite the hostile CF gut, there is clear evidence of probiotic uptake and a reduction in intestinal inflammation (fecal M2PK) from baseline to 12 months. To date, the PEARL-CF study is the only study on probiotics in people with CF to include a HC cohort for comparison and a post-intervention period. This unique design provides further insights into the utility of probiotic supplementation in CwCF.

A recent Cochrane review on probiotics for people with CF identified that probiotics over a 4 to 12 month timeframe may reduce pulmonary exacerbations slightly (low-quality evidence)^21^. The current study identified that probiotic supplementation halved the rates of *H. influenzae* detection in clinically indicated respiratory pathogen tests. The reduction of *H. influenzae* detection rates in the probiotic cohort is likely a result of an effect on the gut-lung axis (GLA)^22,23^, however the exact mechanisms remain unclear. Furthermore, it is unclear if this reflects a reduction in *H. influenzae* lower respiratory tract infections and/or colonization of the upper respiratory tract^24^. Probiotics may be modulating the immune response through enhancing mucosal immunity (e.g. increased production of secretory IgA) and/or regulation of cytokine levels.^25-27^ Further mechanistic investigation is required.

Regarding the fecal microbiota results during the intervention period, the microbiota composition changes substantially over time, however the probiotic only altered this trajectory slightly compared to placebo. The lack of overall community changes in alpha diversity and small change in beta diversity are likely due to several factors including: (i) the hostile intestinal environment in CF outweighing the effect of the probiotic; (ii) the opposite trends observed in alpha diversity for CwCF commencing the probiotic before or after 4 years of age (Fig. 2C-D); and (iii) the inter-individual variability in temporal changes to the intestinal microbial community (Fig. 2A). The lack of change in alpha diversity aligns with findings from CFTR modulator studies, which report either similarly no change in or lower fecal alpha diversity^28-30^. Notably, when probiotic supplementation commenced after 4 years of age, fecal bacterial alpha diversity improved during the intervention period and post-intervention periods, shifting considerably towards levels seen in HC (Fig. 2D and 2G). Thus, in the era of CFTR modulators, there may still be a role for probiotics to improve alpha diversity. Regarding CwCF commencing probiotic supplementation before 4 years of age, there was a non-significant trend showing lower fecal bacterial alpha diversity during probiotic supplementation (Fig. 2C). Probiotic supplementation introduced before the maturation of the intestinal microbiota (approximately 4 years of age; Fig. 2A) may potentially disrupt normal developmental colonization processes. *In vitro* studies have previously identified that *Lactobacillus*-secreted soluble factors contributed to probiotics-induced microbiome inhibition^31^. Additionally, in healthy adults, a multi-strain probiotic (predominantly *Lactobacillus* and *Bifidobacterium* strains) administered post antibiotic therapy showed inhibition of the normal recovery of the fecal microbiota^31^. The clinical relevance of disrupting the intestinal microbiota development is unclear and arguably a reduction in *H. influenzae* detection rates would confer a larger clinical benefit.

Fecal calprotectin is commonly used as a non-invasive biomarker for intestinal inflammation, however in young CwCF, fecal M2PK is a more useful marker^32,33^. This is due to the observation that fecal calprotectin is typically elevated in young children (under 4 years of age) and also lower in CwCF than in HC in the first 3-4 years of life^33^. Fecal M2PK on the other hand is elevated across all ages compared to HC^32^. It appears that this is the first probiotic study in CwCF to utilize fecal M2PK as a biomarker of gut inflammation. Probiotics have previously been shown to reduce fecal M2PK in adults with inflammatory bowel disease and a previous restorative proctocolectomy^34^. The current study demonstrated here that a multi-strain probiotic also reduced fecal M2PK from baseline to the end of 12 months, shifting this biomarker slightly towards levels seen in HC. Given M2PK is expressed in rapidly proliferating cells^35^ and raised in the setting of malignancy and inflammatory states^36^, the reduction seen with probiotic administration likely reflects improved intestinal epithelial homeostasis and reduced cell turnover^37^.

Butyrate-producing taxa *A. butyriciproducens* and *Roseburia hominis* were reduced among CwCF who received probiotics during the intervention period and prevalent in 50% and 21% of participants’ fecal samples, respectively. Consistent with these results, fecal butyric acid levels during the intervention period trended towards being lower in the overall probiotic cohort and were significantly lower in those starting probiotic before 4 years of age. Children with CF commencing probiotics before 4 years of age also had a sustained reduction in *F. prausnitzii* during the post-intervention period (a key butyrate producer present in ∼70% of participants post-intervention fecal samples^38,39^). Similarly, fecal butyric acid levels were significantly lower in the probiotic cohort compared to the placebo cohort at the end of the post-intervention period (24 months). Short-chain fatty acids (SCFA) such as butyrate are generally considered beneficial for both gut and lung health^40^. Butyrate can improve gut immune function, improve epithelial barrier function and reduce inflammation in inflammatory bowel disease^41^. Reductions in *R. hominis* and *F. prausnitzi* have been identified in individuals with ulcerative colitis and may play a role in disease pathogenesis^38^. Butyrate-producing *F. prausnitzii* may also potentially confer protection against bacterial pneumonia^42^, however, the clinical implication(s) of this reduction in endogenous SCFA producers require further investigation. In CwCF commencing probiotics before 4 years of age, the current study also noted an increased relative abundance of *C. difficile* compared to placebo controls during the post-intervention period. The relevance of this finding is unclear, as the prevalence was 27% (of participants post-intervention fecal samples), a rate similar to previous reports of CwCF who are asymptomatic^43^.

Compared to previous probiotic randomized controlled trials (RCTs) in people with CF^21^, the PEARL-CF study had the longest intervention period of 12 months, on par with only Bruzzese et al^44^. The PEARL-CF study is the only study in people with CF to include a post-intervention period, thus providing unique insights into post-intervention effects. In terms of probiotic formulation choice, six of the previous RCTs utilized a single *Lactobacillus* strain and six RCTs utilized a multi-strain formulation (predominantly *Lactobacillus* and *Bifidobacterium* strains), with doses ranging from 100 million to 25 billion colony forming units (CFU) per day^21^. For comparison, the current study used a 15 multi-strain probiotic at a dose of 20 to 30 billion CFU per day.

The PEARL-CF study was impacted by the COVID-19 pandemic, particularly limiting the performance of lung function testing. Due to the limited numbers of lung function tests performed, there was insufficient data to enable any meaningful conclusions on the effects on lung function. The drop-out rate during the intervention period was high and likely attributed to the nature of the study design and sample collection. Despite this, the minimum sample size of 26 participants in each cohort was reached, and retention beyond the intervention period remained high. Although participants younger or older than 3 years of age were block-randomized, a blinded analysis of the fecal alpha diversity indices demonstrated the microbiota was established closer to 4 years of age. Therefore, a cut-off of 4 years of age was employed for all subgroup analyses.

In conclusion, CwCF aged 0-6 years consuming a multi-strain probiotic (predominantly *Lactobacillus* and *Bifidobacterium* strains) had almost 50% fewer clinically indicated respiratory pathogen tests positive for *H. influenzae*. If the probiotic commenced prior to 4 years of age, *H. influenzae* was positive in 70% fewer tests. This study identified a potential therapeutic window for microbiota modulation, as probiotic supplementation commenced prior to 4 years of age may inhibit the normal formative and colonization processes, with consequences to butyrate production, both during the intervention and post-intervention periods. Probiotics commenced after 4 years of age improve fecal bacterial alpha diversity during the intervention and post-intervention periods, thus they may still have a role in the era of CFTR modulators. The various effects of probiotic supplementation in CwCF are likely due to several different mechanisms that require further investigation to elicit the long-term potential clinical benefits. This study provides support for the use of a multi-strain probiotic in young CwCF. Future studies exploring specific probiotic strains and mechanistic pathways are warranted.

## Methods

### Trial oversight

The PEARL-CF study was a double-blind, randomized, placebo-controlled trial of multi-strain probiotic for CwCF, conducted at five sites in Australia and New Zealand: Sydney Children’s Hospital Randwick (SCH), Royal Children’s Hospital Melbourne (RCH), Lady Cilento Children’s Hospital Brisbane (LCHC), The Children’s Hospital at Westmead (CHW), and Christchurch Hospital (CH). This study compared 3 cohorts: (1) CwCF taking probiotics, (2) CwCF taking a placebo, and (3) healthy non-CF controls (not on probiotics/placebo) matched to CF subjects for age and gender.

The trial design was registered and with the Australian and New Zealand Clinical Trials Registry (ACTRN12616000797471p) and the Therapeutic Goods Administration of Australia (CT-2016-CTN-03911-V1). The protocol conformed to the SPIRIT guidelines^45^ and the study was conducted in compliance with Declaration of Helsinki and Good Clinical Practice Guidelines. An external Data and Safety Monitoring Committee (DSMC) reviewed adverse events and study progress.

The trial was funded by the Cystic Fibrosis Foundation, the Australian National Health and Medical Research Council (2020/GNT1194358), Sydney Children’s Hospital Foundation, and the Thrasher Research Fund. Evolution Health Pty Ltd. provided funding support and manufactured, packaged and delivered the probiotic and placebo preparations to all study sites. The funders had no influence on the design or conduct of the trial and were not involved in data collection or analysis, in the writing of the manuscript or in the decision to submit it for publication.

### Study population

Children aged 0–6 years with a confirmed diagnosis of CF (sweat chloride ≥60 mmol/L and/or two CF-causing mutations) were eligible^46^. This study excluded those with prior major intestinal surgery, short bowel syndrome, or any probiotic use in the preceding three months. Concomitant therapies (pancreatic enzymes, standard vitamins, antibiotics) were permitted as clinically indicated.

Healthy controls aged 0–6 years old without CF, other chronic gastrointestinal disorders or any other major disease were eligible. Healthy controls consuming probiotics or with a history of prematurity or low birth weight <2.5kg were not eligible. Healthy controls were matched for age and gender to participants with CF.

Children with CF were recruited from the CF clinics at their respective hospital. Healthy controls were recruited from SCH and the Royal Hospital for Women (RHW).

### Trial procedures

Children with CF were recruited then randomized to a trial arm (Extended Data Figure 13). Participants were provided with a probiotic or placebo and followed for 12 months (intervention period). Participants then ceased the probiotic or placebo and were followed for a further 12 months (post-intervention period). All participants’ families provided written informed consent prior to any study procedures. Stool samples, surveys and clinical data were collected 2-monthly during the intervention period and 3-monthly in the post-intervention period. A time interval range of 4 weeks for each data collection time point was allowed.

Stool samples were collected in a sterile specimen jar using a Feces Catcher (Abbexa Ltd, Cambridge, UK). Samples were transported in a cooler bag (with a −18°C ice pack) to the local hospital laboratory within 24 hours of collection, aliquoted and stored at −80°C. All samples were subsequently transferred to SCH for analysis.

The Qualtrics Online Survey Software Tool (https://www.qualtrics.com/au/) was used to record demographic, clinical and quality of life data at each study timepoint.

### Randomization and blinding

All subjects with CF were randomized to receive either a probiotic supplement or placebo for 12 months with 1:1 allocation using block randomization (blocks of 2, 4 and 6).

Randomization was stratified by site and age group: 0 to 3 years and >3 to 6 years. This age stratification was chosen to determine if there was a therapeutic window prior to the usual establishment of the gut microbiota in healthy controls, which is approximately 3 years of age^7^. The randomization code was computer generated using https://www.sealedenvelope.com and distributed to the Clinical Pharmacists at each site by an independent Pediatric Research Manager from the University of New South Wales. Subject eligibility and recruitment was performed by a study investigator who was blinded to the randomization code and allocation. Allocation was performed by the Clinical Pharmacists at each site, and they had no role in determining subject eligibility or recruitment. Allocation was concealed from both participants and study investigators. The probiotic and placebo were provided to participants in identical 60g glass bottles, differentiated only by a unique study code, ‘Study Group A’ or ‘Study Group B.’ Both preparations were soluble white powders with similar appearance, taste and smell. The Clinical Pharmacists at each site were informed by Evolution Health Pty Ltd regarding the probiotic and placebo allocation if required to address any serious adverse events, however unplanned unblinding was not required at any stage.

Participants, care providers and investigators were all blinded to the randomization throughout the duration of the intervention and post-intervention periods. Healthy controls did not receive a probiotic or placebo.

### Intervention and control

The probiotic powder contained 15-strains, with a combination of *Lactobacillus*, *Bifidobacterium* and *Streptococcus* species (Extended Data Table 1) totaling 10.2 billion colony forming units (CFU) per 1g of powder. This formulation was chosen as no single strain was known to be the most beneficial in CF, and the formulation was similar to a commercially available off-the-shelf product. The placebo composed of maltodextrin and microcrystalline cellulose (MCC) at a ratio of 19:1. The probiotic and placebo powders were soluble and recommended to be given in products such as milk or apple puree. Both the probiotic and placebo could be kept at room temperature.

Children aged 0 to 3 years were administered 2g once daily (∼20.4 billion CFU/day for the probiotic group), whilst children aged ≥3 years were administered 3g once daily (∼30.6 billion CFU/day for the probiotic group). Parents/caregivers were provided with a 2-month supply of the probiotic or placebo powder to maintain integrity of the study products, optimize adherence to the study protocol (data and sample collection) and monitor administration.

Healthy control participants were not given a probiotic or placebo.

### Outcomes

All outcome measures in the PEARL-CF Study were analyzed temporally and cross-sectionally. The group of healthy non-CF control children were included to evaluate the extent to which the intervention restores “normality.”

The primary outcomes include the fecal microbial profiles measured by: (i) microbial alpha diversity measuring using richness at a species level; (ii) microbial alpha diversity measured using Shannon’s diversity index; (iii) microbial beta diversity measured using Bray-Curtis dissimilarity and Aitchison distance^47^; and (iv) individual species that contribute to differences in fecal microbial profiles.

Genomic DNA was extracted from stool samples using the DNeasy PowerSoil Kit (Qiagen, Hilden, Germany) following the manufacturer’s protocol. Approximately 200mg of homogenized stool was transferred to the provided PowerBead tubes containing lysis buffer and mechanical disruption with bead beating was performed to ensure efficient lysis of both gram-positive and gram-negative bacteria. DNA yield and purity were assessed using a NanoDrop 2000 spectrophotometer. Metagenomic libraries were prepared using the Illumina protocol and sequenced on the NovaSeq X Plus platform (Illumina, San Diego, USA) at the Ramaciotti Center for Genomics (UNSW, Sydney, Australia). Sequencing adaptors and low-quality reads (Phred quality ≥ 20) were removed using fastp v0.20.0^48^. Additional polyG tail trimming and base correction by overlap analysis were performed with fastp using the options --trim_poly_g and --correction, respectively. Trimmed reads were mapped to the human genome (GRCh38.p14) with bowtie2 v2.5.2^49^, and unmapped paired-end reads were extracted using samtools v1.19.2^50^. Non-host cleaned reads were analyzed for taxonomic profiling. Taxonomic profiling of bacterial communities was conducted using MetaPhlAn4, following alignment to the MetaPhlAn4 database^51^. The secondary outcomes include: (i) number of pulmonary infections with CF-associated pathogens; (ii) number of hospitalizations for pulmonary exacerbations (defined using Fuch’s criteria^52^); (iii) number of therapeutic antibiotic courses; (iv) anthropometrics measured using height, weight and body mass index z-scores; (v) intestinal inflammation measured by fecal calprotectin (EK-CAL Bühlmann Laboratories; Extended Methods 1); (vi) intestinal inflammation measured by fecal M2PK (Schebo™ TuM2-PK; Extended Methods 1); and (vii) health-related quality of life (HRQOL) measured using age-appropriate PedsQL questionnaires (Extended Methods 2).^53-55^

The tertiary outcomes included optional lung function testing offered to study participants (Extended Methods 3) - using sedated infant lung function (0 to 2 years) or awake multiple breath washout testing (2 to 7 years).

Included in this manuscript is a preliminary analysis of fecal short chain fatty acids (SCFA). Fecal SCFA quantification was performed with liquid chromatography mass spectrometry on the the QExactive (Thermo Scientific, Sydney, Australia) Mass Spectrometer (Extended Methods 4).

### Statistical Analysis

All analyses were conducted according to a modified intention-to-treat principle, with participants being included after providing an initial baseline stool sample and commencing with the probiotic or placebo. Regarding missing data from stool samples for the primary outcomes and fecal biomarkers (calprotectin and M2PK): (i) the last result was carried forward to 12 months if there was a single stool sample collected post commencement of the probiotic/placebo, and (ii) the last result was carried forward to 24 months if there was a single stool sample collected in the post-intervention period.

All analyses were performed using R Statistical Software (v4.5.1; R Core Team 2021) and all outcome measures were analyzed cross-sectionally and temporally. Descriptive statistics were calculated for outcome parameters according to normality of distribution. Categorical variables were compared using Fisher’s Exact Test. Continuous variables were compared using a Student’s *t*-test or Mann-Whitney U test for parametric and non-parametric data, respectively. Longitudinal analyses were performed using linear mixed-effects models (LMMs) to determine the estimated marginal means and adjusted p-values (lme4 package).^56^ Confounders including age and gender were included as random effects in the LMMs.

Microbial alpha diversity was quantified using observed richness (total number of species) and the Shannon-Weaver diversity index. Microbial beta diversity was calculated using Bray-Curtis Dissimilarity and Aitchison distance^57,58^. Principle coordinates analysis (PCoA) was performed to visualize overall community structure. The between-group differences in beta diversity assessed using PERMANOVA (adonis2 package^59^; 999 permutations), and homogeneity of multivariate dispersion verified using betadisper. Differential abundance analysis between cohorts was determined using the MaAsLin 2 (Microbiome Multivariable Associations with Linear Models).^60^ A two-sided P-value of <0.05 was considered statistically significant. Multiple testing correction was applied using the Benjamini–Hochberg procedure, and adjusted p-values (denoted as q-values) reported. All data visualization was performed using the ggplot2 package.^61^

### Sample size calculation

A minimum required sample size of 26 participants on the probiotic, 26 participants on the placebo and 52 healthy controls provided 80% power with a type 1 error probability of 0.05 (two-tailed) to detect an increase in fecal bacterial richness by 75% of the difference between cwCF and healthy controls by the end of 12 months. Estimates of fecal bacterial richness were taken from our previous study on gut microbiota in children with CF and healthy controls (Extended Methods 5).^13^ Assuming a total refusal and drop-out rate of 20%, we aimed to recruit 32 subjects for each CF cohort and 64 healthy controls (matched for age and gender). The sample size was calculated using GLIMMPSE 2.0.0 (https://glimmpse.samplesizeshop.org/<u>#/</u>).

### Ethics and inclusion

The study protocol was approved by the SCH Network Ethics Committee (HREC/16/SCHN/167) and all local institutional review boards. We recruited participants to this study regardless of gender, religion, ethnicity or political views.

### Reporting summary

Further information on the research design is available in the Nature Portfolio Reporting Summary linked to this article.

## Data availability

To adhere to General Data Protection Regulation (https://gdpr-info.eu/), data will not be uploaded to a repository in advance of publication due to the potential for subject identification. Anonymized individual participant data are available upon request from the corresponding author, subject to approval from trial steering groups and data sharing and processing agreements. The timeframe for responding to data requests from the authors is within 1 month.

## Code availability

R code is available for this trial upon request form the corresponding author.

## Acknowledgements

The authors thank the parents and families of our children with CF and our healthy participants, in particular Prof Rebecca Edwards for assisting with the design of this study. This work was supported by the: (i) Cystic Fibrosis Foundation; (ii) CYO was funded by the National Health and Medical Research Council Australia Investigator Grant Fellowship (2020/GNT1194358); (iii) Sydney Children’s Hospital Foundation (iv) Thrasher Research Fund; and (v) Evolution Health Pty Ltd. The authors thank Ms Daniela Arturi for assisting with the probiotic and placebo supply from Evolution Health Pty Ltd.

The authors thank: (i) A/Prof Nancy Briggs, Dr John Widger, Ms Tamarah Katz and Prof Kei Lui for assisting with the protocol development; (ii) Ms Samantha McFedries for providing the randomization code (School of Women’s and Children’s Health UNSW Sydney); (iii) Dr Rishi Bolia (RCH); Ms Samantha Forbes (CHW), Ms Karen McKay (CHW), Ms Ms Andrea Kench (CHW), Dr Shaun Ho (CH), Ms Andreja Fucek (RHW) for participant recruitment and coordination; (iv) Ms Joanne O’Brien (SCH), Ms Donna Legge (RCH), Ms Violet Ford (CHW), Ms Anita Champion (QCH) and Ms Stacey Perret (CH) for dispensing the probiotic and placebo preparations from hospital pharmacies; (v) Dr Louisa Owens, Dr Brendan Kennedy, and Dr Laura Fawcett for performing lung function testing; and (vi) Prof David Ziegler, A/Prof Sean Kennedy and Dr Kylie-Ann Mallitt for sitting on the data safety monitoring board.

## Author contributions

Funding for this study was obtained by CYO, MJC, AJ, TT and A/Prof Nancy Briggs. CYO and MJC were responsible for study conceptualization. CYO, MJC, AJ and TT were responsible for study design, methodology and data analysis plans. CYO and MJC coordinated production and delivery of the study probiotic and placebo. CYO, MJC and J. Halim maintained oversight of study coordination. MJC, J. Halim, RC, NJ, LA, JC, JW, SJM, M. Corbett, AD, MO, SR, HS and CW recruited and/or coordinated participants. LP performed lung function testing. MJC and J. Halim maintained the study database. JvD, UP, J. Halim and J. Hudson performed the processing of fecal samples for analysis. MJC, JvD, UP, KC and CF curated and performed the metagenomic bioinformatic analyses and data interpretation. MJC curated and performed the clinical data analysis and interpretation. MJC drafted and wrote the final manuscript. All authors critically reviewed and approved the manuscript.

## Additional information

**Extended data** are available for this paper at:

## Supplementary information

The online version contains supplementary material available at:

**Correspondence and requests for materials** should be addressed to Dr Michael Coffey

## Extended Data Tables & Figures

**Extended Data Table 1.** Probiotic formulation.

| Strains | CFUs |
| --- | --- |
| *Bifidobacterium lactis* BI-04 | 3 Billion |
| *Lactobacillus rhamnosus* GG | 2 Billion |
| *Lactobacillus paracasei* Lpc-37 | 1 Billion |
| *Lactobacillus plantarum* Lp-115 | 1 Billion |
| *Lactobacillus rhamnosus* HN001 | 500 Million |
| *Lactobacillus rhamnosus* Lr32 | 500 Million |
| *Bifidobacterium animalis* HN019 | 500 Million |
| *Lactobacillus salivarius* subsp. *salivarius* Ls-33 | 400 Million |
| *Streptococcus thermophiles* St21 | 400 Million |
| *Bifidobacterium breve* Bb-03 | 200 Million |
| *Lactobacillus gasseri* Lg 36 | 200 Million |
| *Bifidobacterium longum* BI-05 | 180 Million |
| *Bifidobacterium infantis* BI-26 | 100 Million |
| *Lactobacillus reuteri* 1e1 | 100 Million |
| *Lactobacillus bulgaricus* Lb-64 | 100 Million |
| **Excipient:** Litesse Ultra (Polydextrose) | 950 mg |
| **Total (15 Strains) CFU per 1g:** | 10.2 Billion |

CFU, colony forming units

**Extended Data Figure 1.**
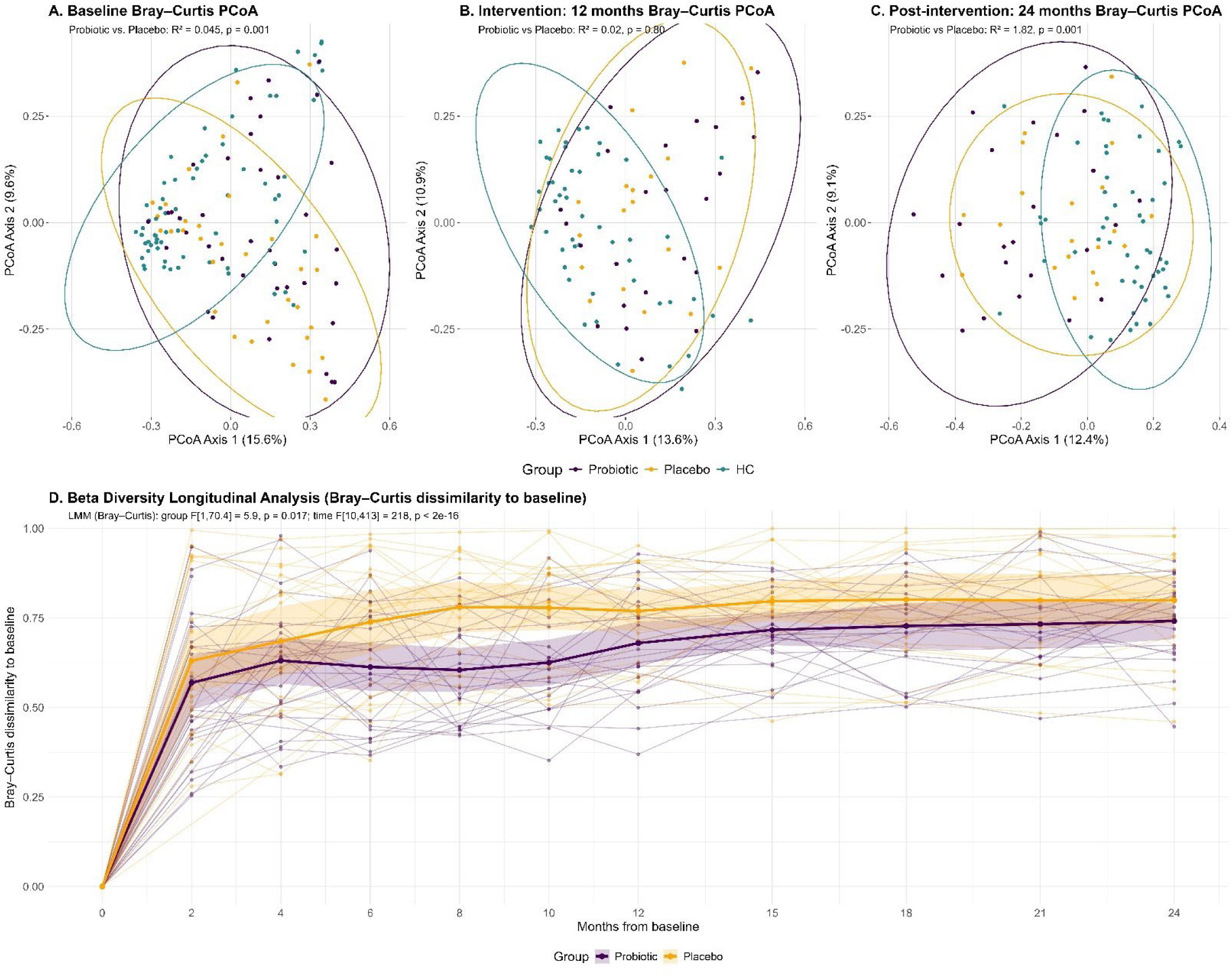
Fecal bacterial beta diversity (Bray Curtis Dissimilarity) Principal Coordinates Analysis (PCoA) of Bray–Curtis dissimilarity based on species-level relative abundances (with 95% confidence ellipses) at baseline (**A**), end of the 12 month intervention period (**B**), and end of the 24 month post-intervention period (**C**). In graphs A-C, PERMANOVA results are for probiotic vs. placebo analyses (not including healthy controls). **Fig. D** presents a longitudinal analysis of beta diversity trends from baseline to 24 months for all CwCF. Lines show individual trajectories with overlaid group means and 95% confidence intervals. Linear mixed-effects models were used to analyse treatment group and time effects.

**Extended Data Table 2.** Differentially abundant fecal bacterial species during the intervention period.

| **Fecal bacterial species** | **Coefficient (SE)** | **Prevalence** | **P-value** | **Q-value** |
| --- | --- | --- | --- | --- |
| ↑ *Lactiplantibacillus plantarum* * | 5.0 (0.7) | 32% | 6.9E-10 | 2.8E-07 |
| ↑ *Lacticaseibacillus paracasei* * | 3.8 (0.7) | 45% | 3.6E-07 | 0.00004 |
| ↑ *Bifidobacterium breve* * | 3.8 (1.3) | 64% | 0.005 | 0.07 |
| ↑ *Ligilactobacillus salivarius* * | 3.1 (0.5) | 24% | 1.2E-07 | 0.00002 |
| ↑ *Lacticaseibacillus rhamnosus* * | 3.1 (0.8) | 57% | 0.0003 | 0.008 |
| ↑ *Bifidobacterium animalis* * | 2.6 (0.9) | 61% | 0.007 | 0.08 |
| ↑ *Limosilactobacillus reuteri* * | 2.4 (0.4) | 24% | 5.8E-08 | 0.00001 |
| ↑ *Lactobacillus gasseri* * | 2 (0.5) | 26% | 0.0002 | 0.008 |
| ↓ *Enterococcus SGB6173* | -1.1 (0.5) | 39% | 0.04 | 0.24 |
| ↓ *Roseburia hominis* | -1.4 (0.6) | 21% | 0.03 | 0.22 |
| ↓ *Bifidobacterium dentium* | -1.4 (0.7) | 15% | 0.034 | 0.23 |
| ↓ *Coprococcus comes* | -1.5 (0.6) | 26% | 0.02 | 0.17 |
| ↓ *Roseburia intestinalis* | -1.5 (0.7) | 26% | 0.042 | 0.24 |
| ↓ *Mediterraneibacter glycyrrhizinilyticus* | -1.7 (0.8) | 39% | 0.037 | 0.23 |
| ↓ *Agathobaculum butyriciproducens* | -2.2 (1) | 50% | 0.03 | 0.22 |

\* Bacterial species included in the probiotic formulation. ↑ and ↓ indicates if the bacterial species was increased or decreased in the probiotic cohort relative to the placebo cohort. SE, standard error. Prevalence is the number of participants (combined probiotic and placebo cohorts) that have the bacterial species detected in their stool sample.

**Extended Data Figure 2.**
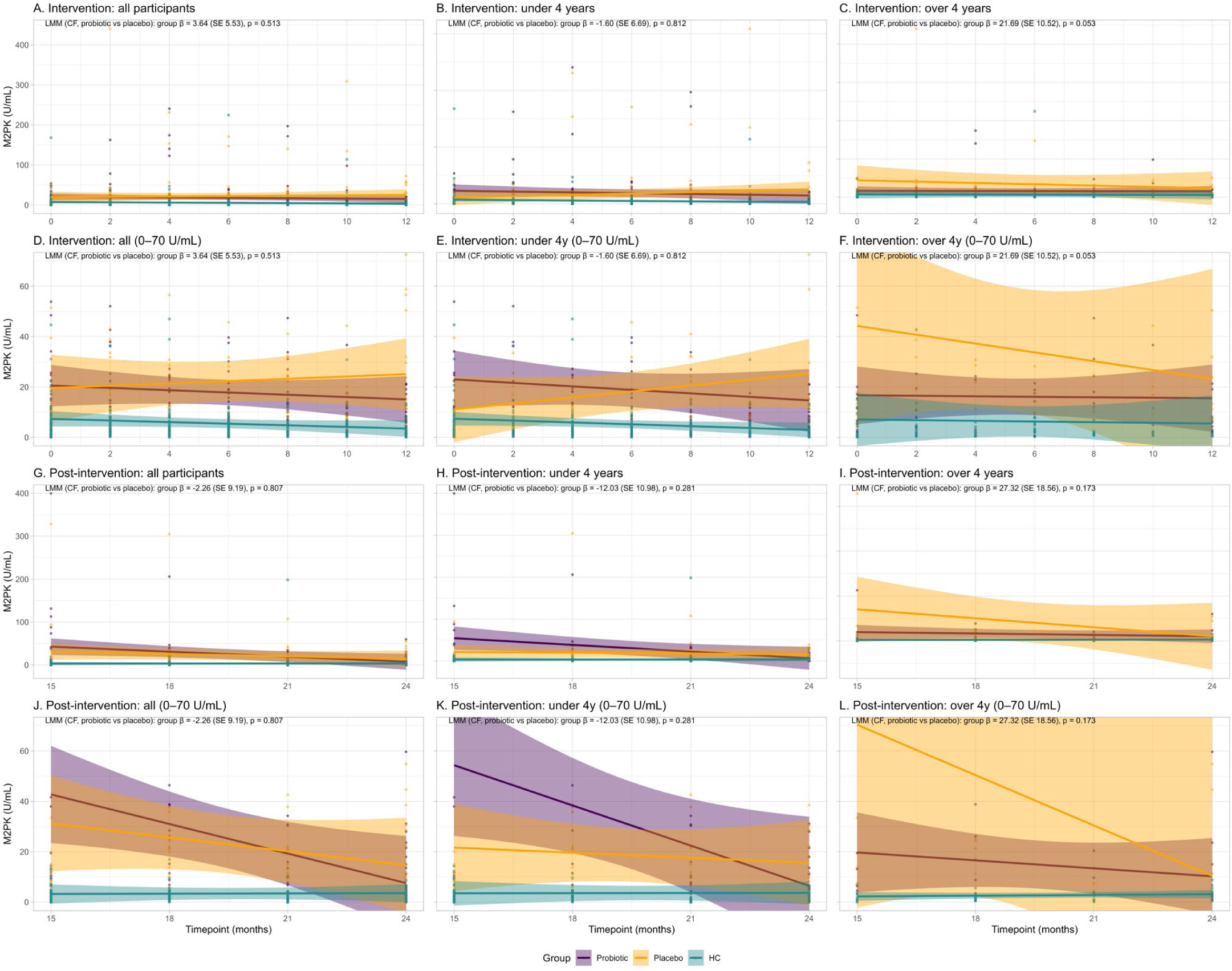
Fecal M2-pyruvate kinase (M2PK) Panels **A–C** show M2PK levels across the intervention period for all participants (**A**), children aged <4 years (**B**), and ≥4 years (**C**). Panels **D–F** present zoomed views restricted to 0–70 U/mL. Panels **G–I** depict post-intervention trajectories using the full scale, while panels **J–L** show zoomed post-intervention results (0–70 U/mL). Data points and regression lines represent individual measurements and linear model estimates by intervention group (Probiotic, Placebo, Healthy Controls), with shaded bands indicating 95% confidence intervals. Healthy controls are included for visual comparison only. Linear mixed-effects models were performed on CF participants only (excluding healthy controls), adjusted for age and time, with model annotations displaying the estimate (β), standard error (SE), and *p*-value for the probiotic vs placebo group effect.

**Extended Data Figure 3.**
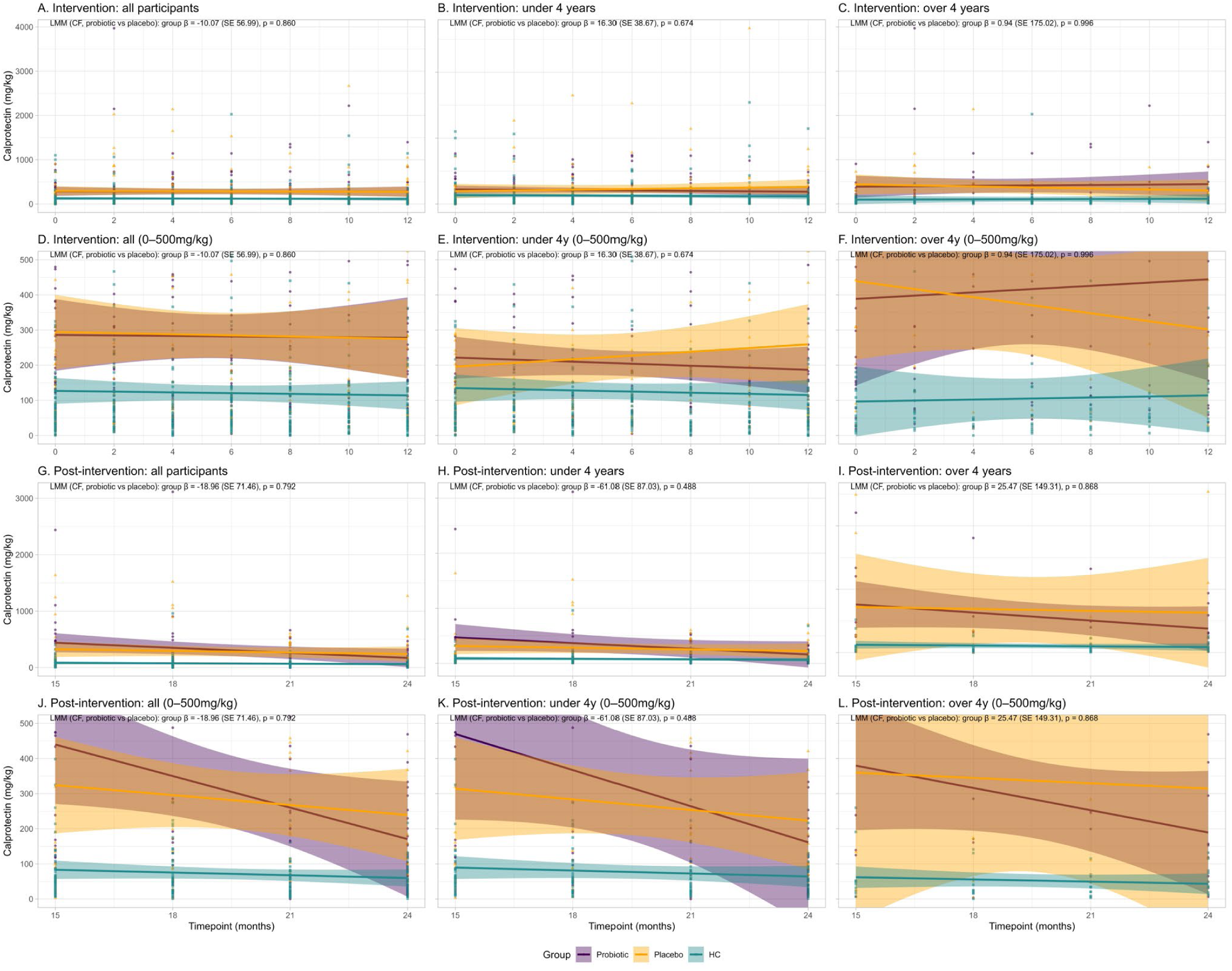
Fecal calprotectin. Panels **A–C** show fecal calprotectin levels across the intervention period for all participants (**A**), children aged <4 years (**B**), and ≥4 years (**C**). Panels **D–F** present zoomed views restricted to 0–500 mg/kg. Panels **G–I** depict post-intervention trajectories using the full scale, while panels **J–L** show zoomed post-intervention results (0–500 mg/kg). Data points and regression lines represent individual measurements and linear model estimates by intervention group (Probiotic, Placebo, Healthy Controls), with shaded bands indicating 95% confidence intervals. Healthy controls are included for visual comparison only. Linear mixed-effects models were performed on CF participants only (excluding healthy controls), adjusted for age and time, with model annotations displaying the estimate (β), standard error (SE), and *p*-value for the probiotic vs placebo group effect.

**Extended Data Table 3.** Lung Function Testing Numbers.

|  | **Probiotic** | **Placebo** |
| --- | --- | --- |
| **Infant Lung Function** |  |  |
| Baseline Test | 2 | 2 |
| 12-month Test | 1 | 0 |
| **Pre-school Lung Function (MBW)** |  |  |
| Baseline Test | 7 | 4 |
| 12-month Test | 0 | 3 |
| **School-age Spirometry** |  |  |
| Baseline Test | 8 | 2 |
| 12-month Test | 2 | 0 |

**Extended Data Figure 4.**
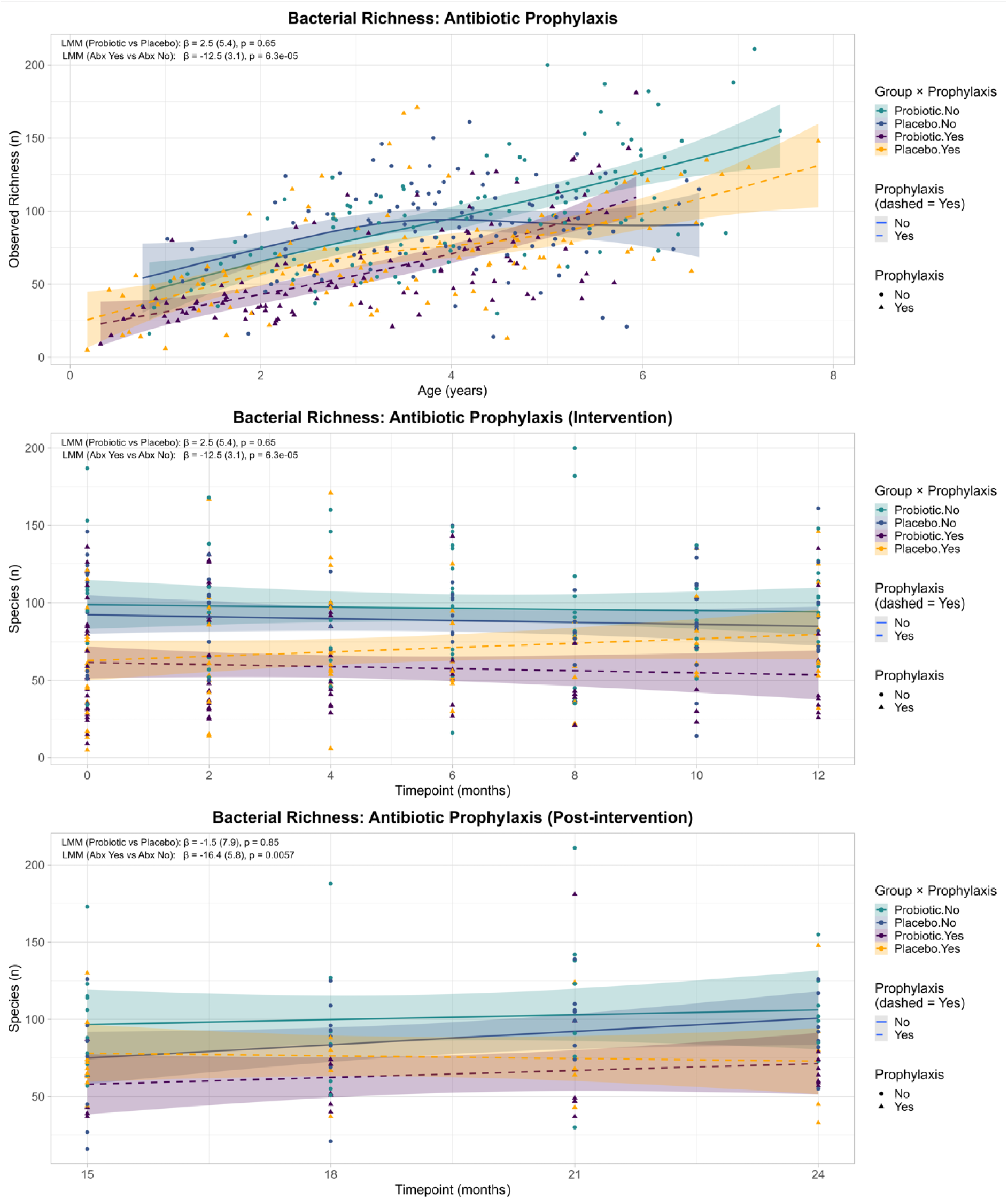
Antibiotic Prophylaxis Sensitivity Analysis of Alpha Diversity. (A) Cross-sectional analysis of observed species richness by age, stratified by treatment group and current prophylactic antibiotic use. (B) Longitudinal trends during the 12-month intervention period. (C) Post-intervention follow-up (15–24 months). Lines indicate model-based estimates with 95% confidence intervals. Linear mixed-effects models were adjusted for treatment group, age, and timepoint. Prophylactic antibiotic use was associated with reduced bacterial richness, while probiotic vs placebo effects remained consistent.

**Extended Data Figure 5.**
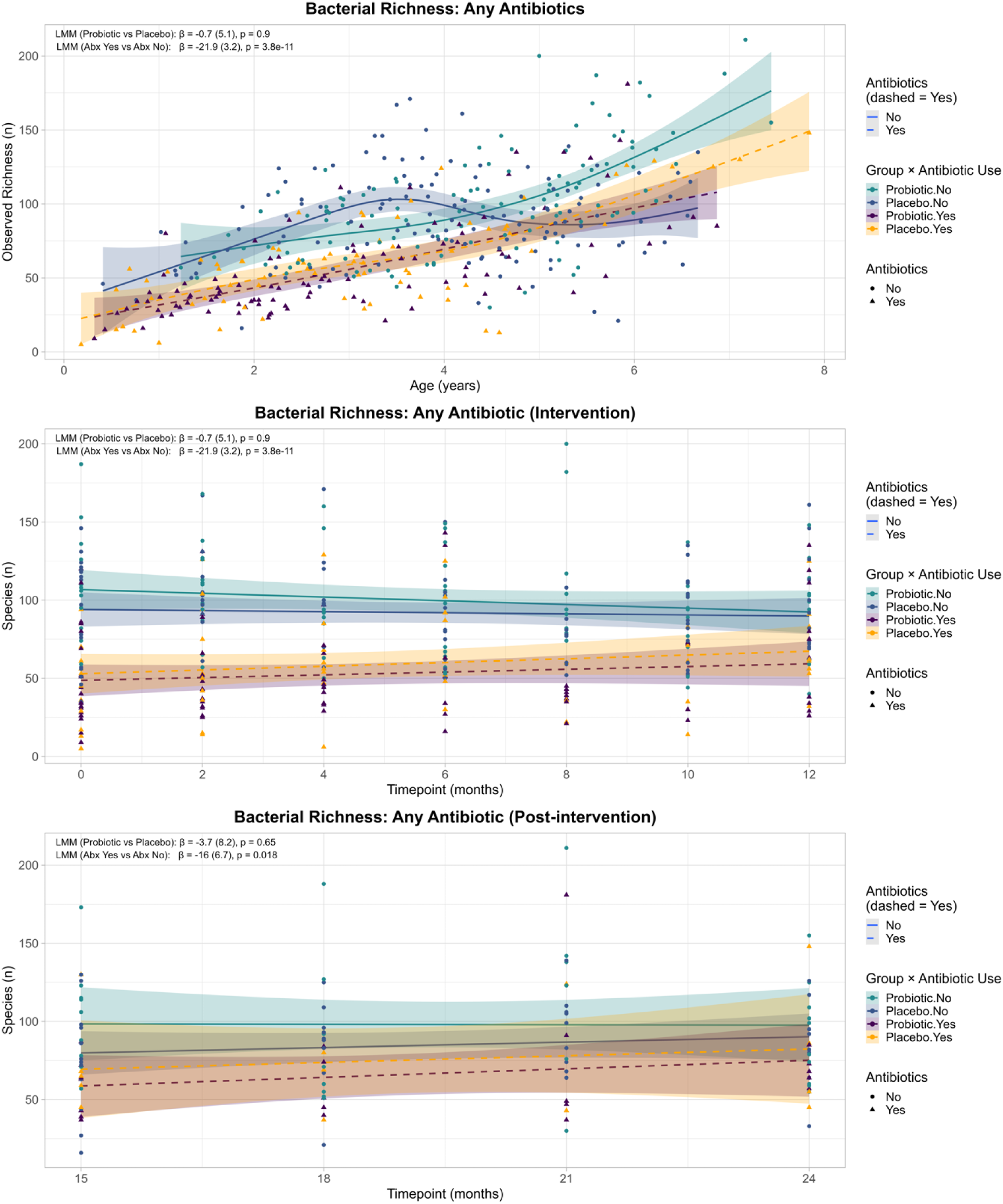
Any Antibiotic Use Sensitivity Analysis of Alpha Diversity. (A) Observed species richness by age across treatment and current antibiotic use categories. (B) Longitudinal changes over the 12-month intervention period. (C) Post-intervention period (15–24 months). Lines represent model estimates with 95% confidence intervals. Linear mixed-effects models accounted for treatment group, age, and timepoint. Antibiotic use was associated with lower richness, without evidence of interaction with treatment allocation.

**Extended Data Figure 6.**
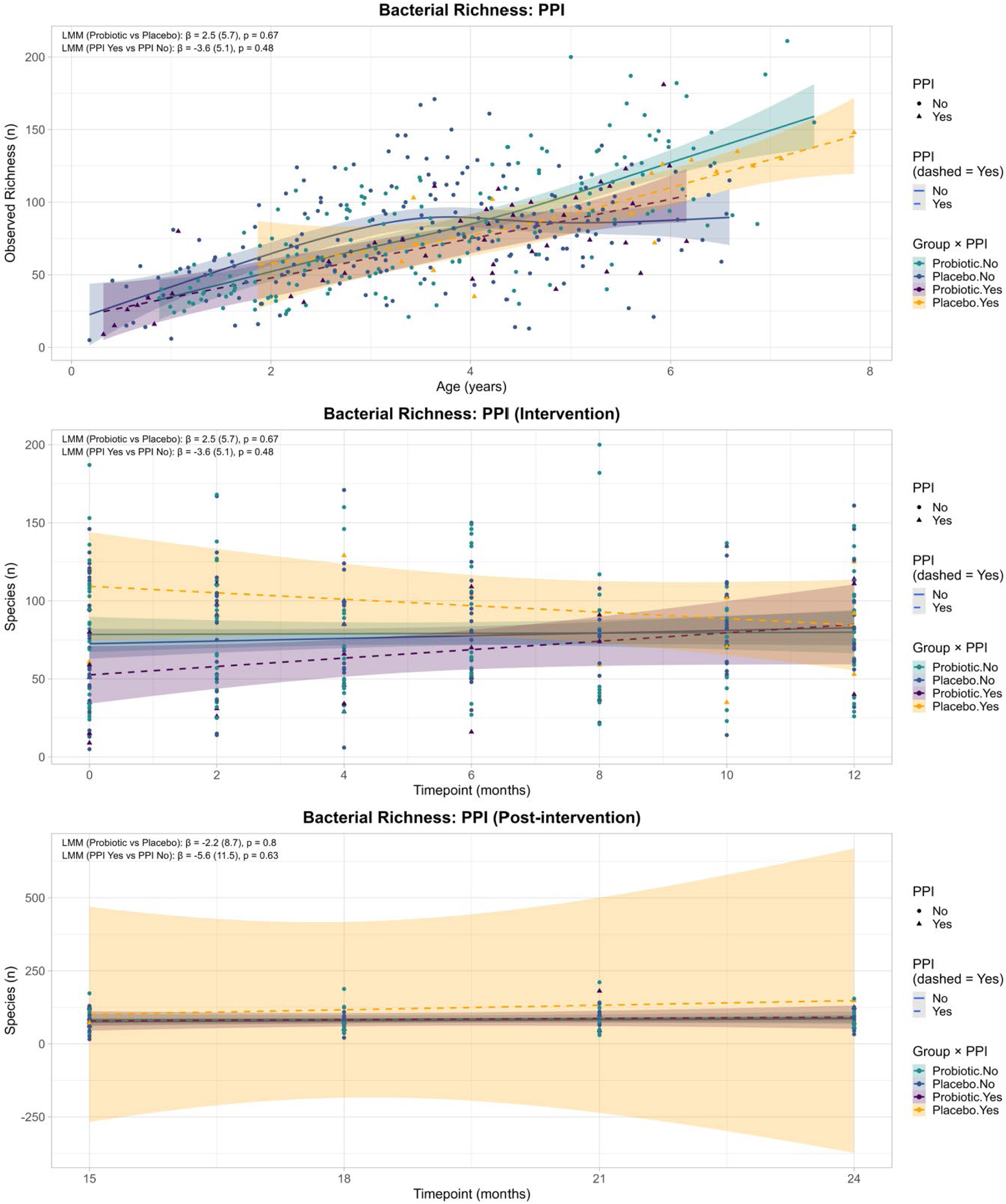
Proton Pump Inhibitor Use Sensitivity Analysis of Alpha Diversity. (A) Observed species richness by age across treatment and current PPI use categories. (B) Longitudinal changes over the 12-month intervention period. (C) Post-intervention period (15–24 months). Lines represent model estimates with 95% confidence intervals. Linear mixed-effects models accounted for treatment group, age and timepoint. PPI use was not significantly associated with richness, with no evidence of interaction with treatment allocation.

**Extended Data Figure 7.**
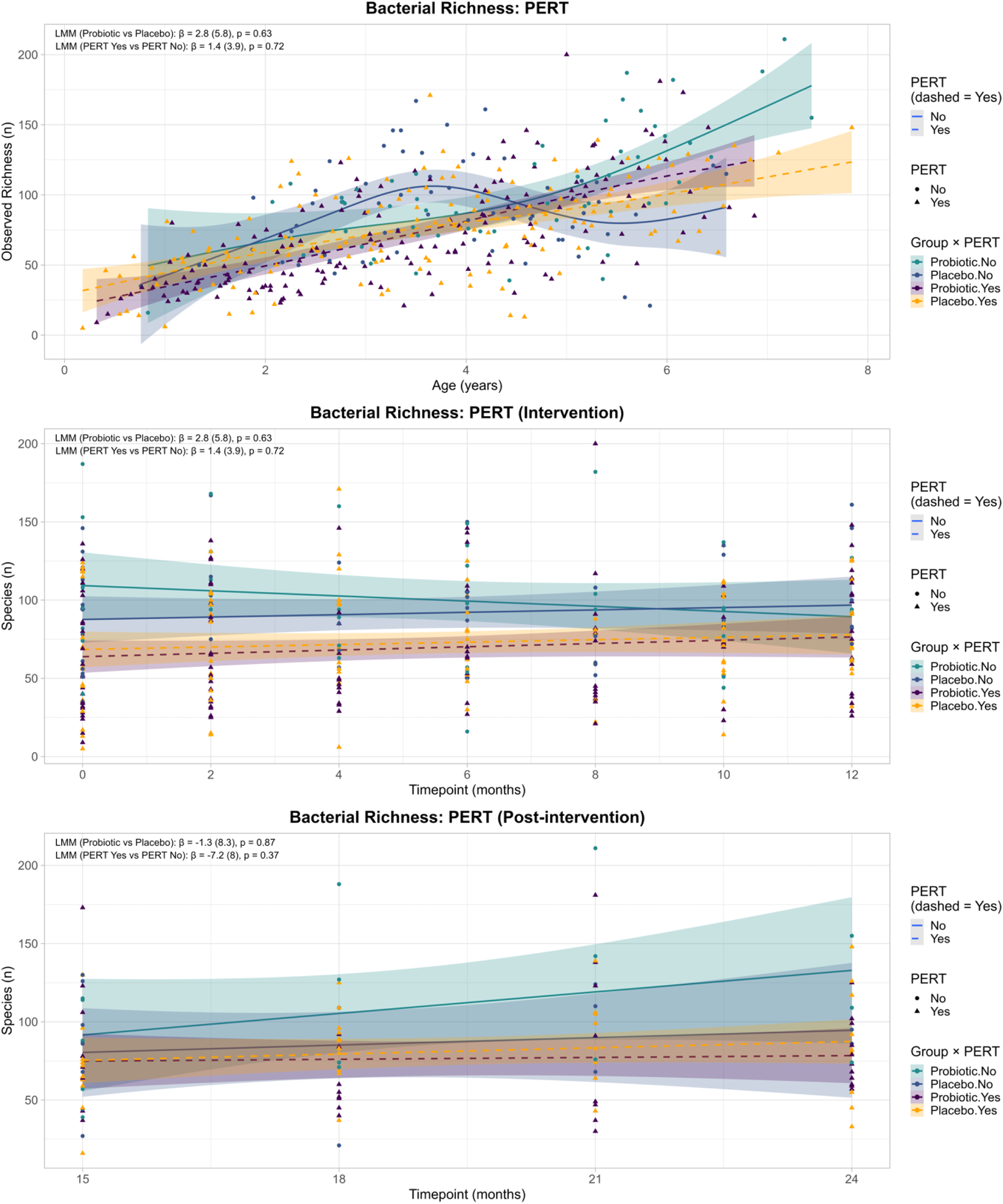
Pancreatic Enzyme Replacement Therapy (PERT) Use Sensitivity Analysis of Alpha Diversity. (A) Observed species richness by age stratified by treatment and PERT use. (B) Longitudinal trends during the 12-month intervention period. (C) Post-intervention follow-up (15–24 months). Lines show model estimates with 95% confidence intervals. Linear mixed-effects models adjusted for treatment group, age and timepoint. PERT use showed no significant association with richness, and treatment effects were unchanged.

**Extended Data Figure 8.**
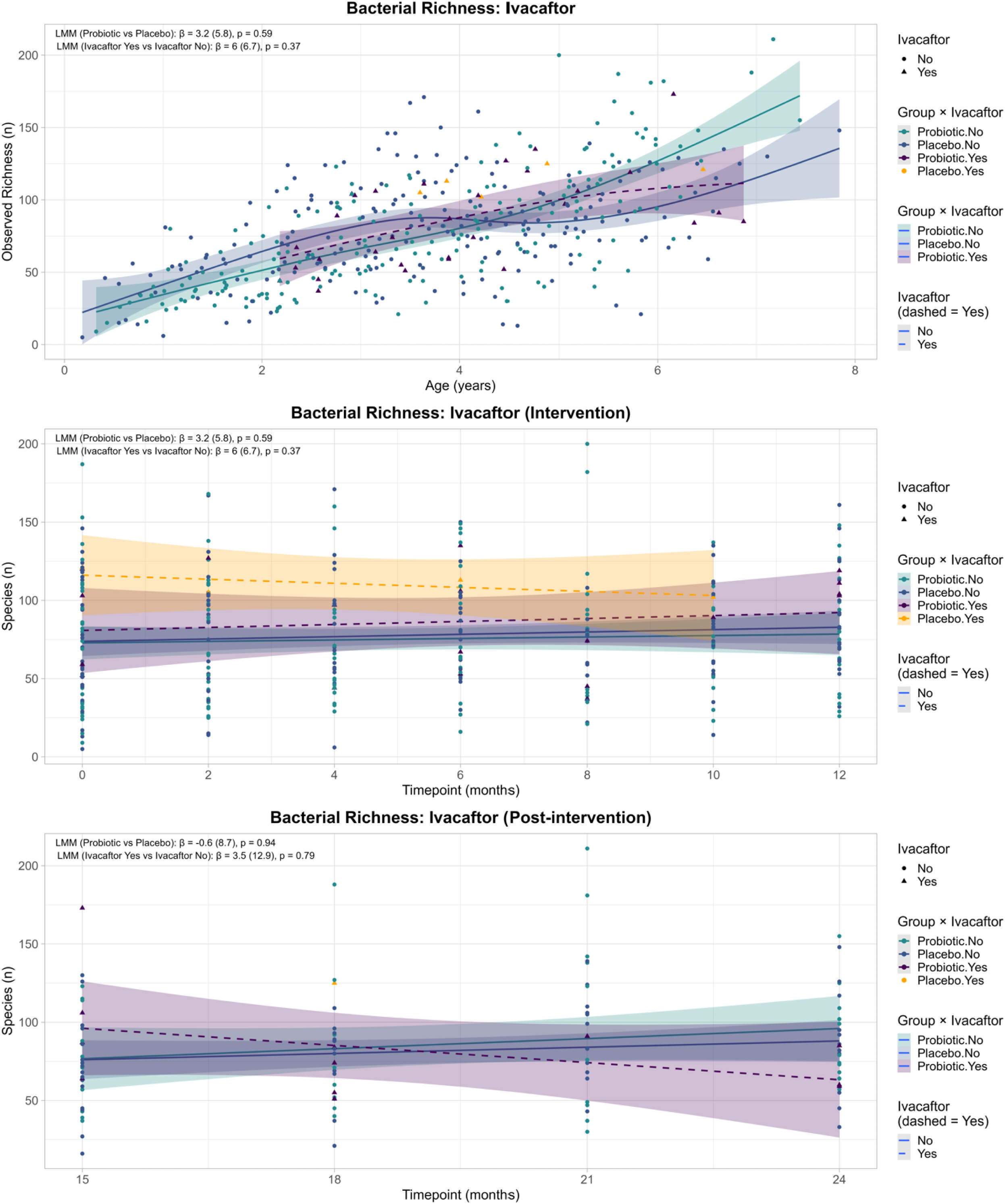
Ivacaftor Use Sensitivity Analysis of Alpha Diversity. (A) Observed richness by age according to treatment and ivacaftor use. (B) Longitudinal richness during the 12-month intervention. (C) Post-intervention period (15–24 months). Lines indicate model-based trends with 95% confidence intervals. Models were adjusted for treatment group, age and timepoint. Ivacaftor use was not significantly associated with richness, and did not modify the probiotic treatment effect.

**Extended Data Figure 9.**
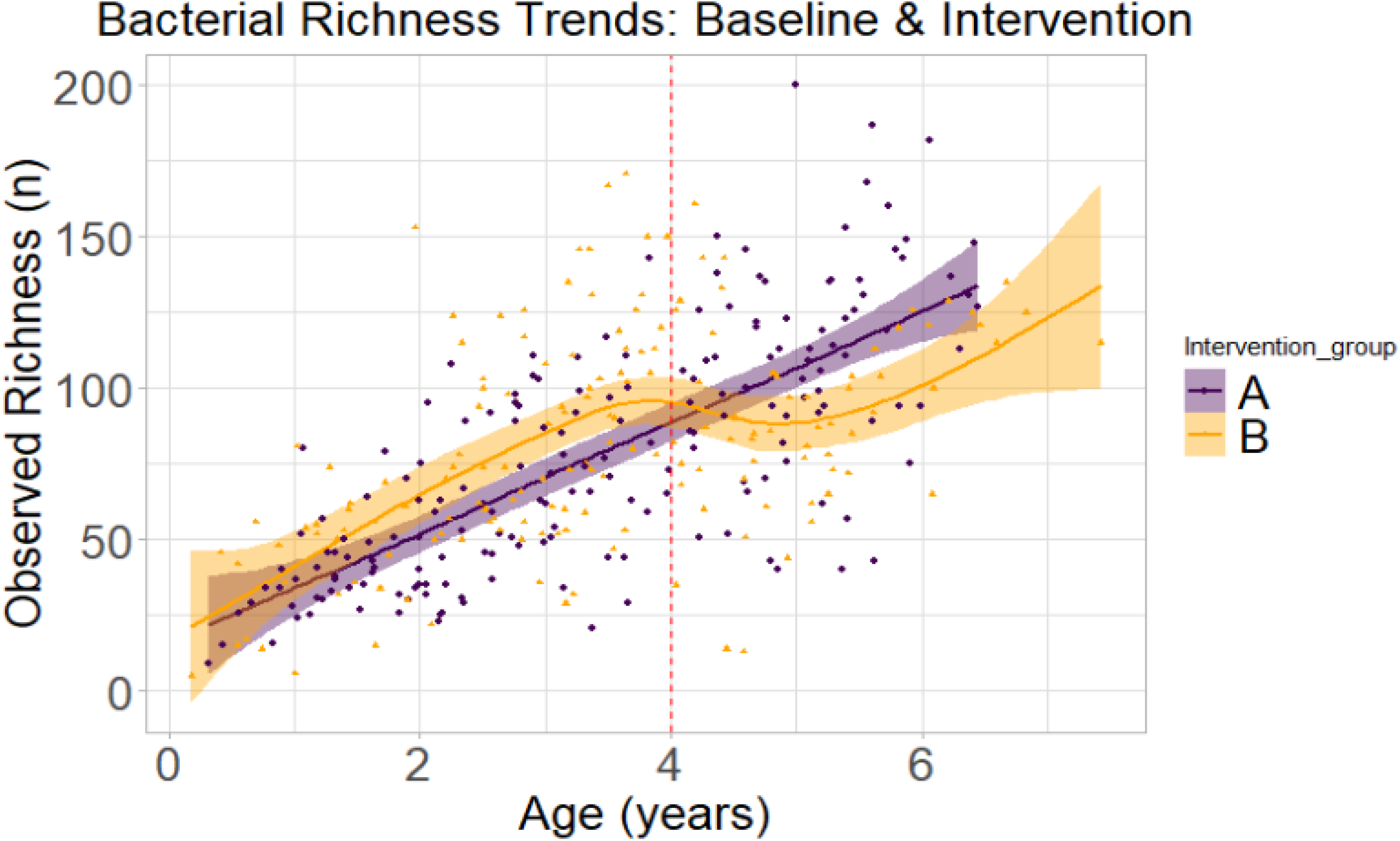
Blinded analysis of fecal alpha diversity (observed richness).

**Extended Data Figure 10.**
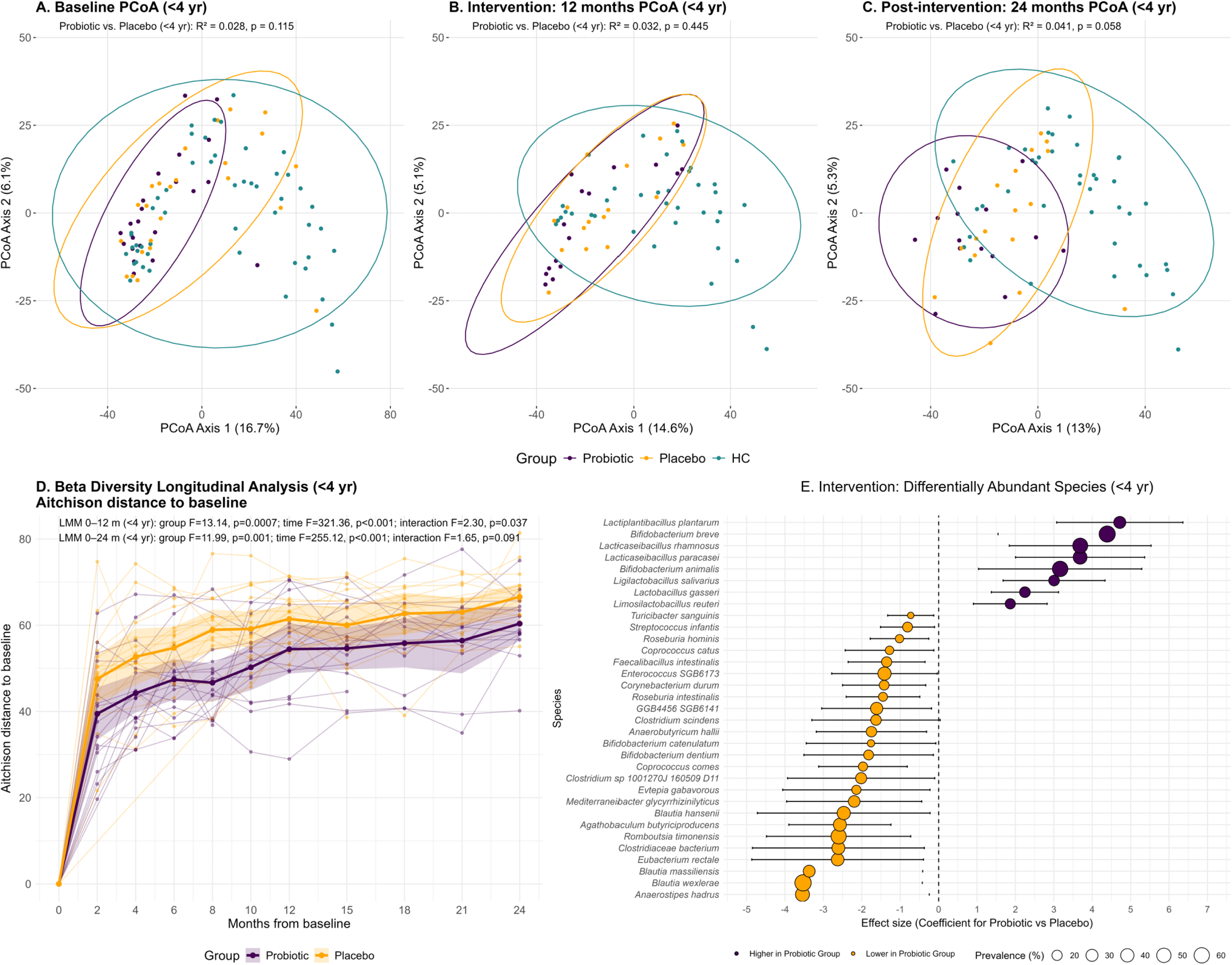
Fecal bacterial beta diversity (Aitchison distance) and differentially abundant fecal bacterial species (during the intervention period) for CwCF commencing the probiotic/placebo prior to 4 years of age. Principal Coordinates Analysis (PCoA) of Bray–Curtis dissimilarity based on species-level relative abundances (with 95% confidence ellipses) at baseline (**A**), end of the 12 month intervention period (**B**), and end of the 24 month post-intervention period (**C**). In graphs A-C, PERMANOVA results are for probiotic vs. placebo analyses (not including healthy controls). **Fig. D** presents a longitudinal analysis of beta diversity trends from baseline to 24 months for all CwCF aged less than 4 years of age at the start of probiotic/placebo supplementation. Lines show individual trajectories with overlaid group means and 95% confidence intervals. Linear mixed-effects models were used to analyse treatment group and time effects.

**Extended Data Table 4.**
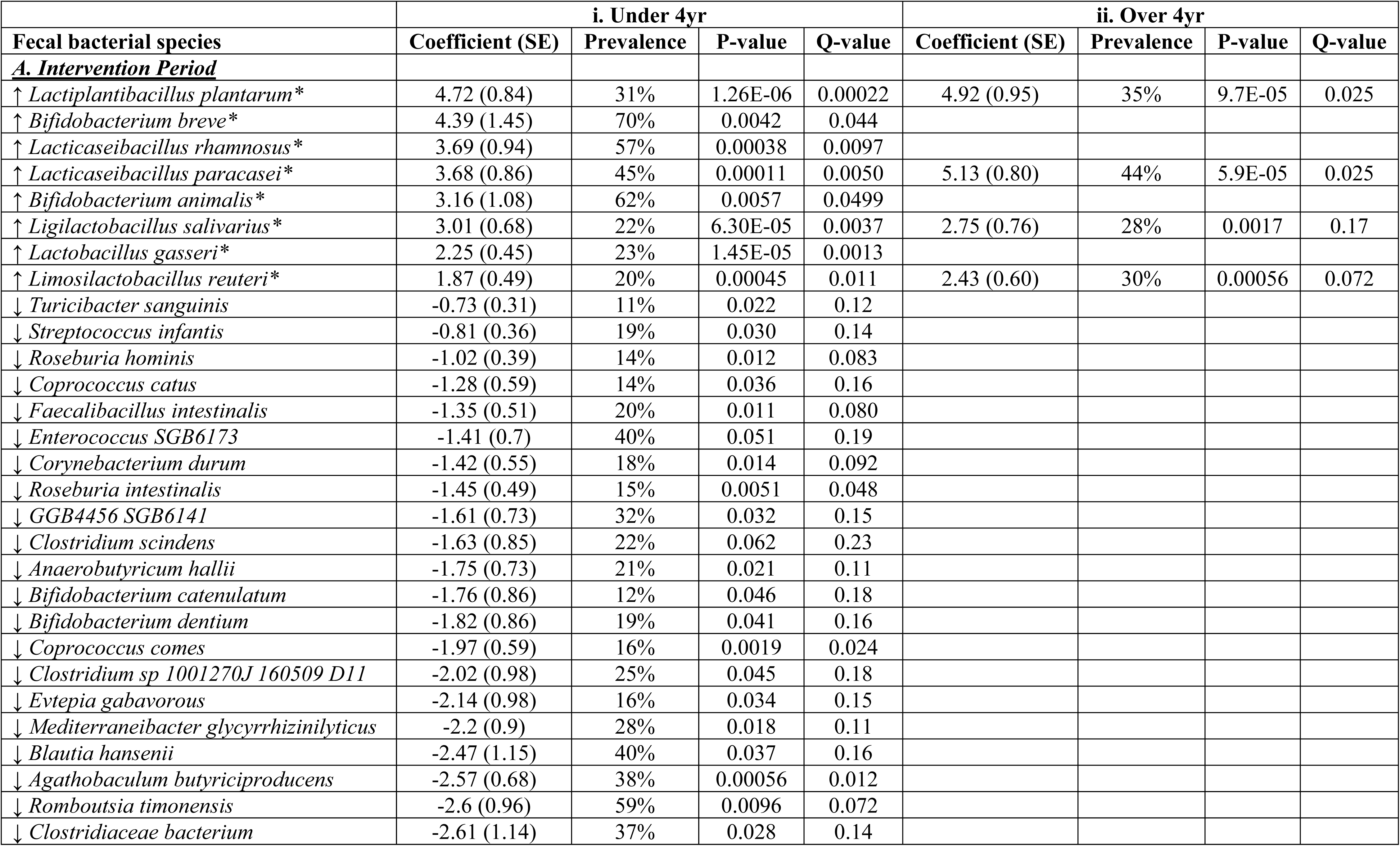
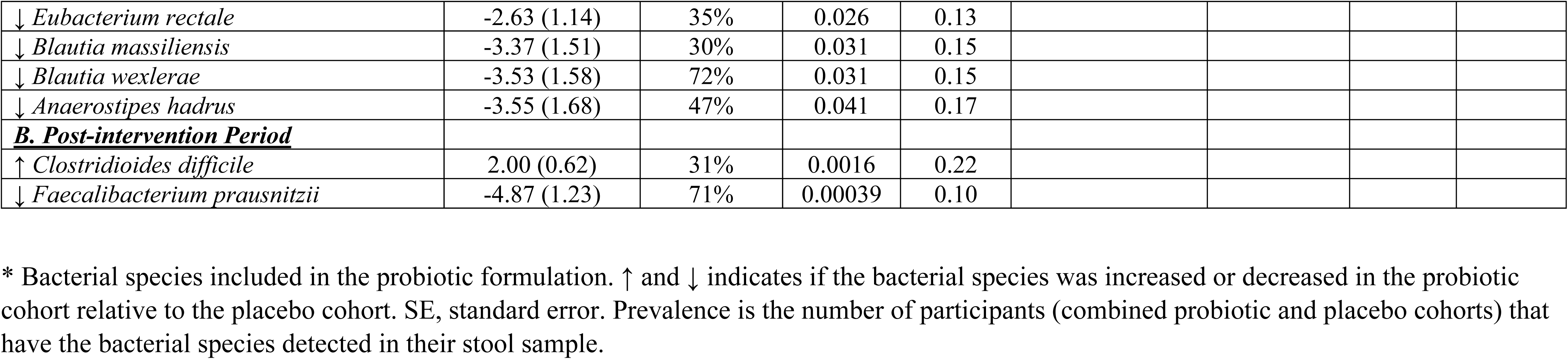
Post hoc analysis of differentially abundant fecal bacterial species during the intervention and post-intervention periods.

**Extended Data Figure 11.**
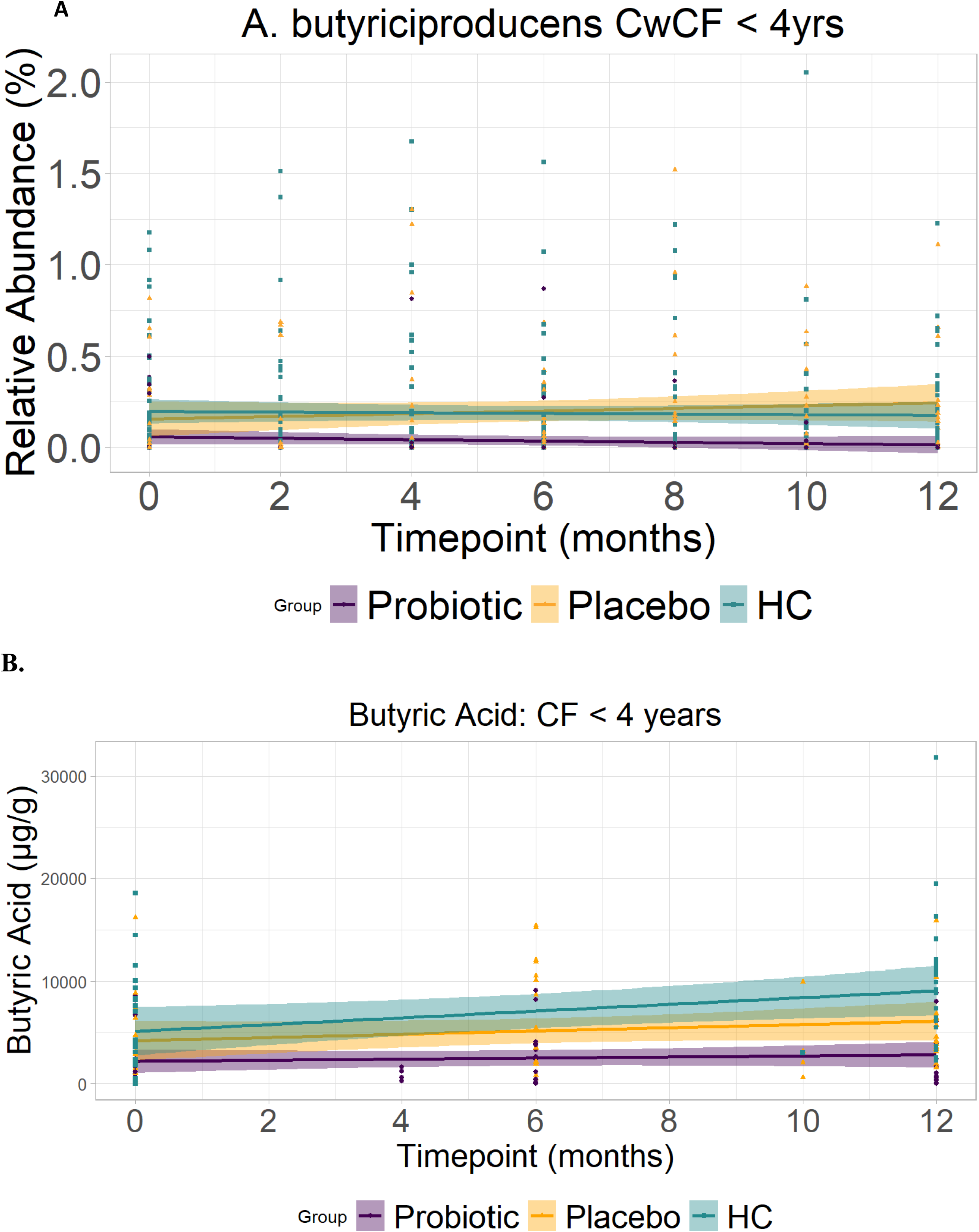
Fecal *A. butyriciproducens* and butyric acid levels during the intervention period in CwCF < 4 years of age.

**Extended Data Figure 12.**
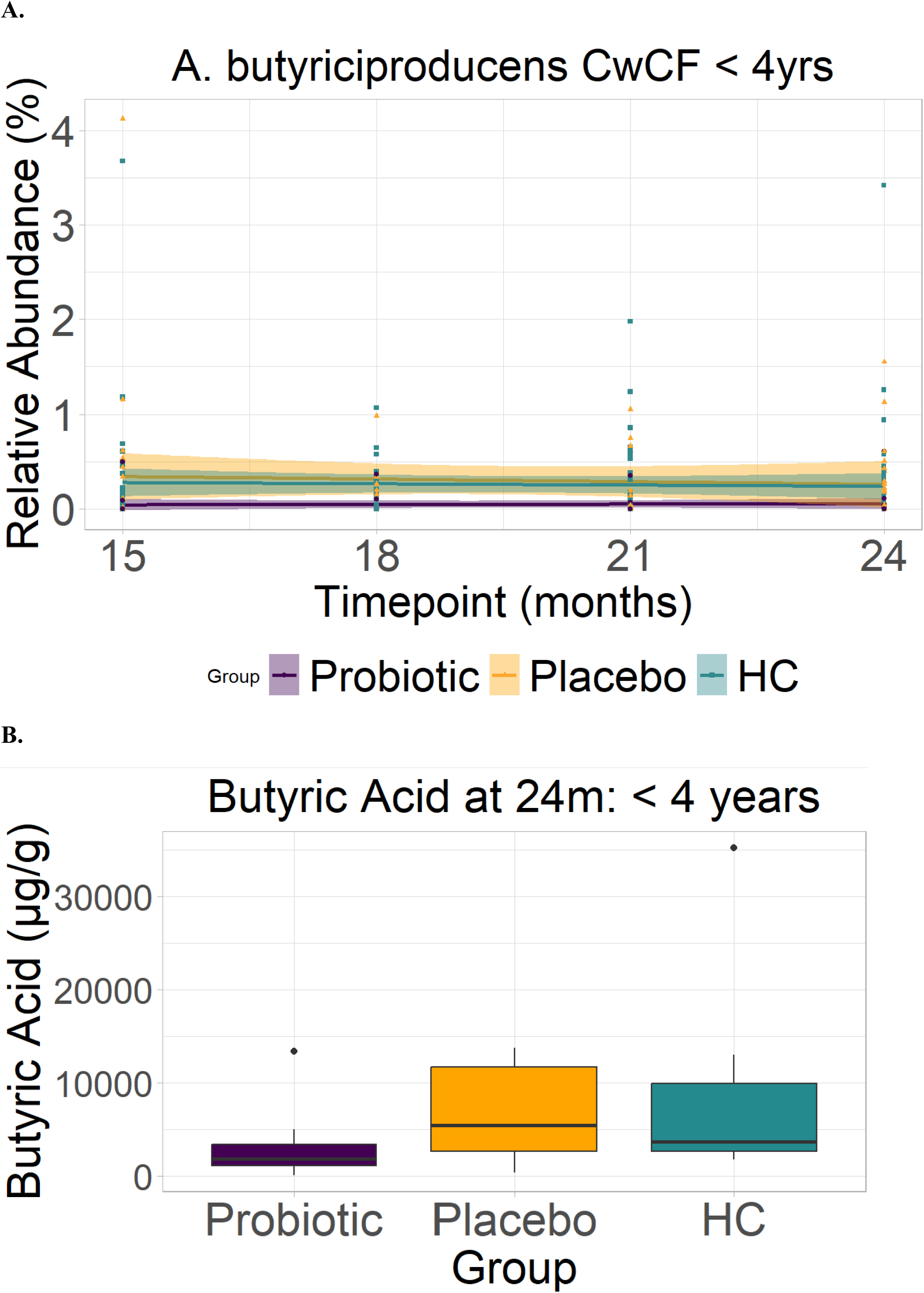
Fecal *A. butyriciproducens* and butyric acid levels during the post-intervention period in CwCF < 4 years of age.

**Extended Data Figure 13.**
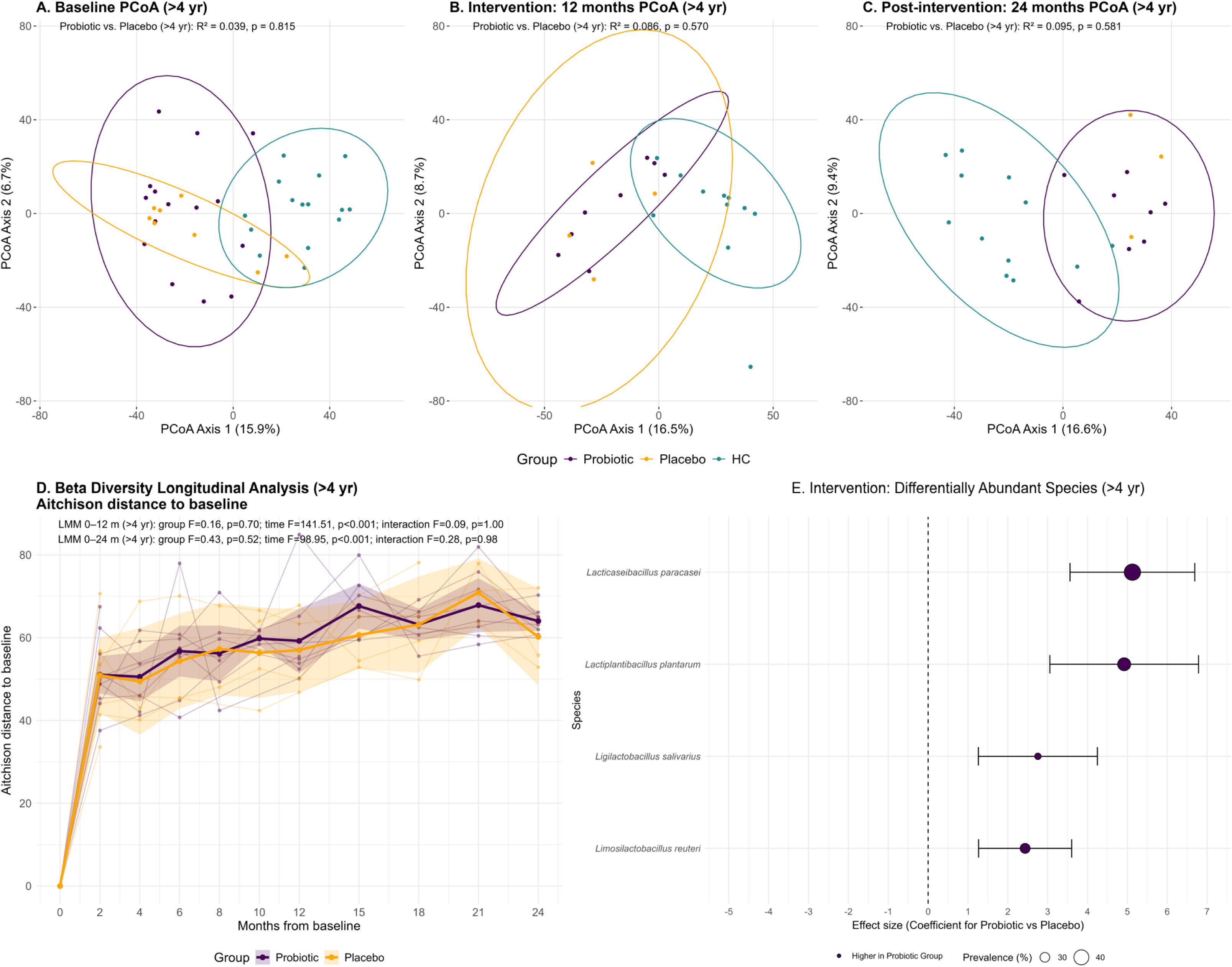
Fecal bacterial beta diversity (Aitchison distance) and differentially abundant fecal bacterial species (during the intervention period) for CwCF commencing the probiotic/placebo after 4 years of age. Principal Coordinates Analysis (PCoA) of Bray–Curtis dissimilarity based on species-level relative abundances (with 95% confidence ellipses) at baseline (**A**), end of the 12 month intervention period (**B**), and end of the 24 month post-intervention period (**C**). In graphs A-C, PERMANOVA results are for probiotic vs. placebo analyses (not including healthy controls). **Fig. D** presents a longitudinal analysis of beta diversity trends from baseline to 24 months for all CwCF aged 4 years or older at the start of probiotic/placebo supplementation. Lines show individual trajectories with overlaid group means and 95% confidence intervals. Linear mixed-effects models were used to analyse treatment group and time effects.

**Extended Data Figure 14.**
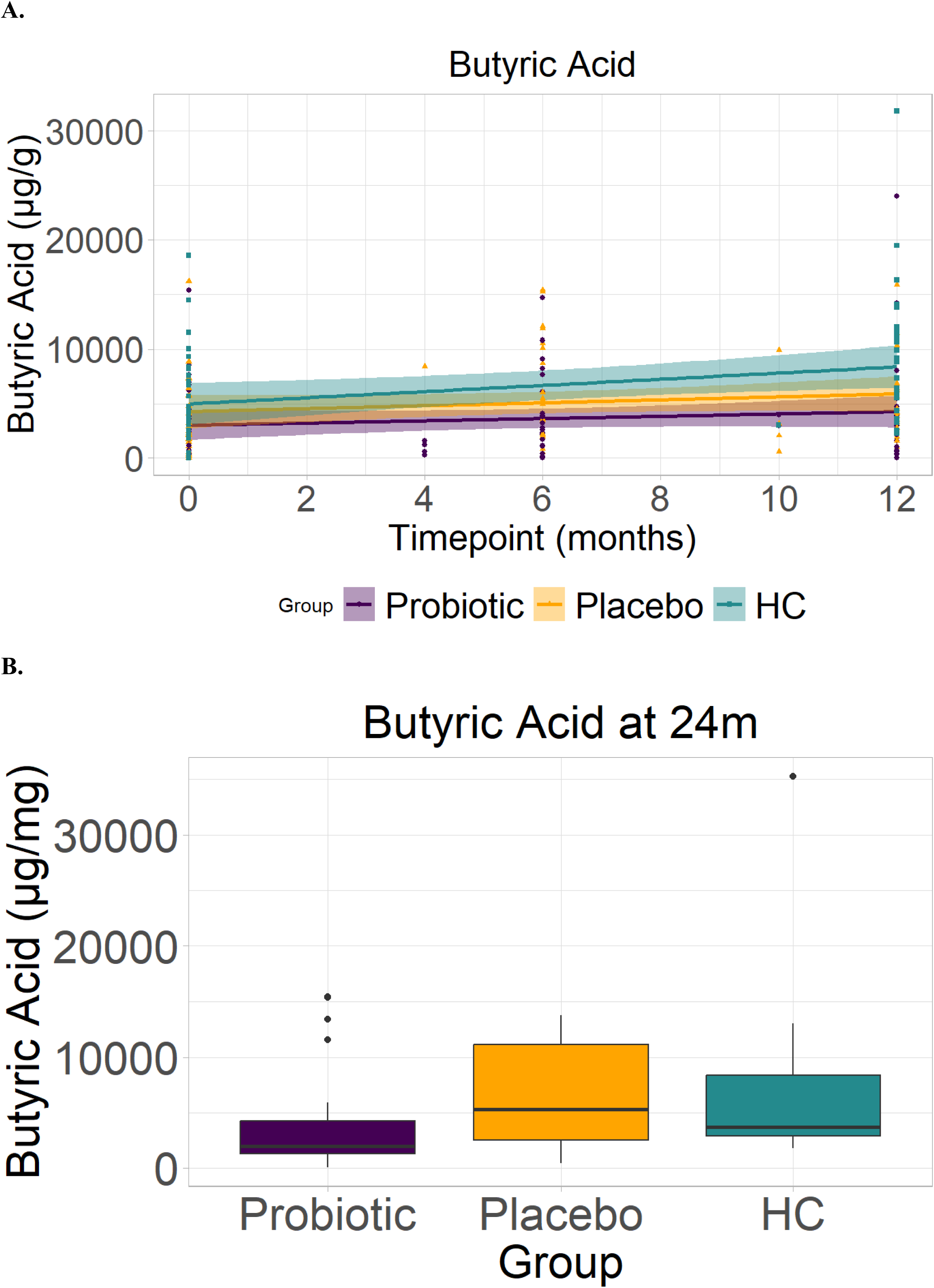
Fecal butyric acid levels during the intervention period (A) and at the end of the post intervention period (B) in all cwCF and HC.

**Extended Data Figure 15.**
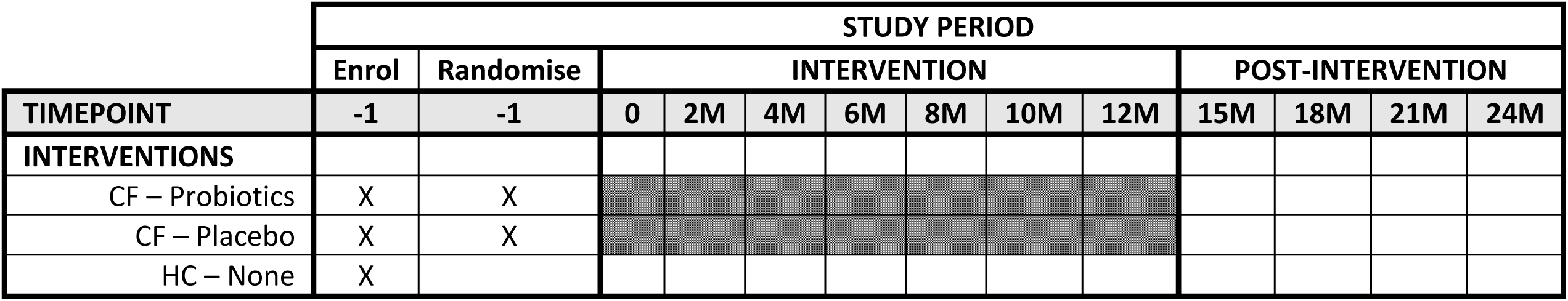
Study protocol and timeline. 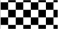 Daily administration of probiotic or placebo; CF, cystic fibrosis, HC, healthy control.

**Extended Methods 1.** Fecal calprotectin and M2 pyruvate kinase (M2PK).

Fecal calprotectin was quantified using a monoclonal enzyme-linked immunosorbent assay (ELISA) EK-CAL Calprotectin ELISA, Bühlmann Laboratories, Switzerland, according to the manufacturer’s protocol. Samples exceeding the upper assay limit (600 µg/g) were diluted and re-analysed to obtain quantitative results.

Fecal M2PK concentrations were measured from stool samples with the Schebo™ TuM2-PK ELISA test kit.

All samples were measured in duplicate, according to the manufacturer’s instructions. Any concentrations outside the detection limit range were further diluted and repeated to ensure accurate calculation of concentrations.

**Extended Methods 2.** Health-related quality of life questionnaires. The following PedsQL questionnaires were used:

- ages 1 to 12 months - Infant Scales v1.0 (Parent report for ages 1-12 months);^1^
- ages 13 to 24 months – Infant Scales v1.0 (Parent report for ages 13-24 months);^1^
- ages 2 to 4 years – Gastrointestinal Symptoms Module v3.0 (Parent report for ages 2-4);^2,3^
- ages 5 to 7 years – Gastrointestinal Symptoms Module v3.0 (Parent report for ages 5-7).^2,3^

**Extended Methods 3.** Optional lung function testing

Multiple breath washout (MBW) testing was performed using the Exhalyzer D device by EcoMedics AG (Switzerland), and used to quantify the lung clearance index (LCI).

Infant lung function testing in children under 2 years of age was performed under conscious sedation with oral chloral hydrate at 100mg/kg, up to a maximum of 1g.^4^ The MBW testing was performed using 4% sulfa hexa-fluride (SF6). Finally, the Raised Volume Rapid Thoraco-abdominal Compression (RV-RTC) test was used to mimic the forced expiratory flow maneuvers of spirometry.^4,5^

Preschool lung function testing was performed in children aged 2 to 7 years. The MBW testing was performed using nitrogen and a washout with 100% oxygen.

**Extended Methods 4.** Fecal short chain fatty acid analysis

SCFA quantification was performed with liquid chromatography mass spectrometry (LC-MS) on the QExactive (Thermo Scientific) Mass Spectrometer using the Agilent C18 1.9 m, 100 x 2.1 mm column. Samples, spiked QC samples, and calibrants were extracted with acetonitrile and derivatised using 3-NPH. Isotopically labelled internal standards for SCFAs were derivatised using 13C6-3-NPH.[41]. Six SCFA derivatives were quantified; acetic (AA), propionic (PA), butyric (BA), valeric (VA), isobutyric (IBA) and isovaleric (IVA) acids. Standard curve dilutions were prepared in 50% acetonitrile and spiked with internal standards. Final concentrations were calculated in μg/g dry weight.

**Extended Methods 5.** Data for sample size calculation

The required sample size for this study was calculated using the following conditions: (i) we assume that patients with CF who take probiotics over 12 months will achieve an improvement in the number of species, such that it increases by 75% of the difference between CF patients and healthy controls by the end of 12 months; (ii) we used estimates of the mean for each measure at 3 time points, 0, 6 and 12 months; (iii) relative group sizes of 1:1:2 for CF-probiotic, CF-placebo and healthy controls respectively; (iv) our estimates are calculated to detect a main effect only of groups; (v) type 1 error probability of 0.05 (two-tailed); (vi) desired power of 0.8; (vii) the GLIMMPSE Software (http://glimmpse.samplesizeshop.org/#/) was used.

The mean fecal bacterial richness (Log10 operational taxonomic units (OTUs)) taken from Nielsen et al., 2016:^6^

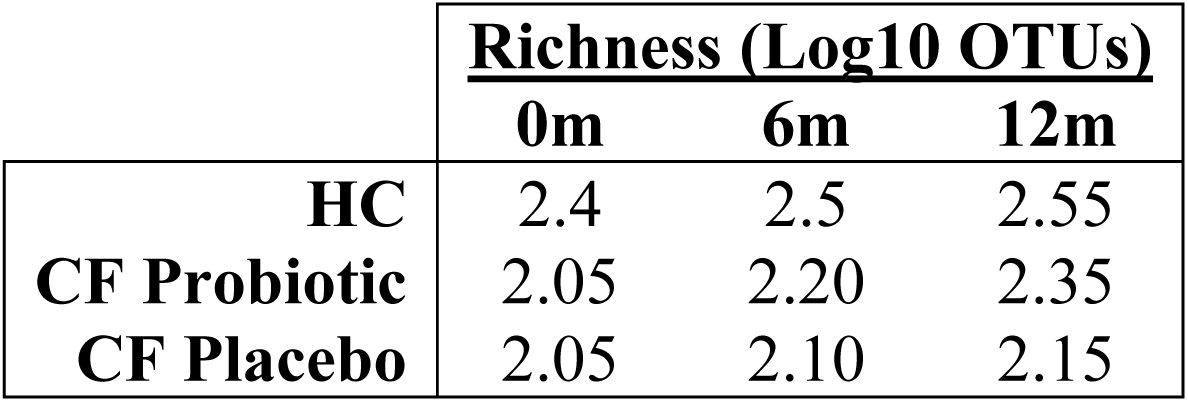

Results from the sample size calculation are presented below:

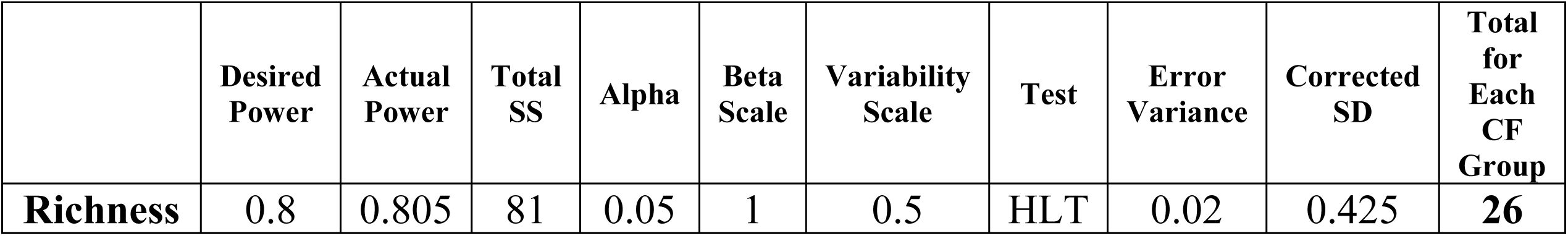

|  | **Desired Power** | **Actual Power** | **Total SS** | **Alpha** | **Beta Scale** | **Variability Scale** | **Test** | **Error Variance** | **Corrected SD** | **Total for Each CF Group** |
| --- | --- | --- | --- | --- | --- | --- | --- | --- | --- | --- |
| **Richness** | 0.8 | 0.805 | 81 | 0.05 | 1 | 0.5 | HLT | 0.02 | 0.425 | **26** |

